# A Tool for Translating Polygenic Scores onto the Absolute Scale Using Summary Statistics

**DOI:** 10.1101/2021.04.16.21255481

**Authors:** Oliver Pain, Alexandra C. Gillett, Jehannine C. Austin, Lasse Folkersen, Cathryn M. Lewis

**Author notes:** **Corresponding author:** Dr. Oliver Pain, Social, Genetic and Developmental Psychiatry Centre, Institute of Psychiatry, Psychology and Neuroscience - PO80, De Crespigny Park, Denmark Hill, London, United Kingdom, SE5 8AF.

## Abstract

**Background:** There is growing interest in the clinical application of polygenic scores as their predictive utility increases for a range of health-related phenotypes. However, providing polygenic score predictions on the absolute scale is an important step for their safe interpretation. Currently, polygenic scores can only be converted to the absolute scale when a validation sample is available, presenting a major limitation in the interpretability and clinical utility of polygenic scores.

**Methods:** We have developed a method to convert polygenic scores to the absolute scale for binary and normally distributed phenotypes. This method uses summary statistics, requiring only the area-under-the-ROC curve (AUC) or variance explained (*R*^2^) by the polygenic score, and the prevalence of binary phenotypes, or mean and standard deviation of normally distributed phenotypes. Polygenic scores are converted using normal distribution theory. Given the AUC/*R*^2^ of polygenic scores may be unknown, we also evaluate two methods (AVENGEME, lassosum) for estimating these values from genome-wide association study (GWAS) summary statistics alone. We validate the absolute risk conversion and AUC/*R*^2^ estimation using data for eight binary and three continuous phenotypes in the UK Biobank sample.

**Results:** When the AUC/*R*^2^ of the polygenic score is known, the observed and estimated absolute values were highly concordant. Across binary phenotypes, the mean absolute difference between the observed and estimated proportion of cases was 5%. For continuous phenotypes, the mean absolute difference between observed and estimated means was <0.3%. Estimates of AUC/*R*^2^ from the lassosum pseudovalidation method were most similar to the observed AUC/*R*^2^ values, though estimated values deviated substantially from the observed for autoimmune disorders.

**Conclusion:** This study enables accurate interpretation of polygenic scores using only summary statistics, providing a useful tool for educational and clinical purposes. Furthermore, we have created interactive webtools implementing the conversion to the absolute scale for binary and normally distributed phenotypes (https://opain.github.io/GenoPred/PRS_to_Abs_tool.html). Several further barriers must be addressed before clinical implementation of polygenic scores, such as ensuring target individuals are well represented by the GWAS sample.

## Introduction

A substantial proportion of individual differences in health and disease are explained by genetic variation (Polderman et al., 2015). Genome-wide association studies (GWAS) have successfully identified thousands of genetic loci associated with a broad range of phenotypes, from anthropometric traits such as height (A. R. Wood et al., 2014), to psychiatric disorders such as major depression (Howard et al., 2019). One application of GWAS results is the estimation of genetic risk/propensity for a given phenotype using polygenic scores. Polygenic scores are calculated as the GWAS effect size-weighted sum of alleles carried by an individual (Choi, Mak, & O’Reilly, 2020). A range of methods exist for processing GWAS summary statistics prior to polygenic scoring to account for the linkage disequilibrium (LD) between variants and improve the predictive utility of polygenic scores (Pain et al., 2020).

Polygenic scores capture only part of the genetic liability to a disease or trait, but the proportion explained increases with larger GWAS sample sizes and with improvements in polygenic scoring methodology. For example, individuals in the top 8% of cardiovascular disease (CAD) polygenic scores have a three-fold increased risk of developing CAD compared to the general population (Khera et al., 2018). The implementation of polygenic scores within a clinical setting are being increasingly discussed and investigated (Lewis & Vassos, 2020; Wray et al., 2021), but several barriers exist before these scores can become an established part of healthcare, including the low variance of risk explained and issues with interpretation.

For correct interpretation of a polygenic score, the score must first be standardised according to an ancestry-matched reference, so that the score is transformed to units of standard deviations from the mean. This standardised score, referred to as a polygenic Z-score, can then be used to calculate the relative risk of disease for an individual, based on their polygenic score. Whilst relative risk estimates are of interest, they are challenging to interpret as they do not consider the predictive utility of the polygenic score or the prevalence of the outcome in the general population. It is well established that differences in risk are most accurately perceived by non-experts when presenting risk on the absolute scale, i.e., the probability an individual will develop the outcome (Gigerenzer, Gaissmaier, Kurz-Milcke, Schwartz, & Woloshin, 2007; Zipkin et al., 2014). The absolute risk conferred by a given relative risk is determined by the predictive utility of the polygenic score and the population prevalence of the phenotype. For example, an individual’s polygenic Z-score for schizophrenia may be 1.96, indicating their polygenic score is higher than 97.5% of an ancestry matched population. However, the absolute risk, or probability of the individual developing schizophrenia, is unknown, as we must account for the predictive utility of the polygenic score and the population prevalence of schizophrenia.

When individual-level data is available for the polygenic score and the outcome of interest, it is possible to calculate the absolute risk conferred by a given polygenic score by measuring the proportion of cases within polygenic scores quantiles. This approach is often used in research to put variance explained estimates into perspective (Khera et al., 2018), and it is also implemented by 23andMe to help interpretation of results by their customers (Furlotte, Kleinman, Smith, & Hinds, 2015). However, individual-level data for the phenotype of interest within a representative sample is often unavailable. Therefore, a summary statistic-based approach, requiring only information describing the predictive utility of the polygenic score and the prevalence of the outcome, would greatly improve the availability of interpretable results from polygenic scores.

Within a homogenous population, polygenic scores are normally distributed due to the central limit theorem, and therefore normal distribution theory can be used to define polygenic scores based on the distribution of the phenotype and predictive utility of the polygenic score. Using this approach, our study develops a summary statistic-based tool for converting polygenic scores into absolute estimates for both binary and normally distributed phenotypes. For binary phenotypes, we calculate absolute risk based on the predictive utility of the polygenic score and the population prevalence of the phenotype. For normally distributed phenotypes, we calculate predictions in absolute terms based on the predictive utility of the polygenic scores and the population mean and standard deviation of the phenotype. In addition, we compare several approaches that estimate the predictive utility of polygenic scores using GWAS summary statistics alone, as this value is often unknown.

Another important factor reported to help accurately interpret differences in risk is the use of simple visual aids (Zipkin et al., 2014). Therefore, this study also develops interactive webtools for converting an individual’s polygenic scores to the absolute scale with corresponding graphics.

## Methods

### UK Biobank (UKB)

UKB is a prospective cohort study that recruited >500,000 individuals aged between 40-69 years across the United Kingdom (Bycroft et al., 2018). The UKB received ethical approval from the North West - Haydock Research Ethics Committee (reference 16/NW/0274).

#### Phenotype data

Eleven UKB phenotypes were analysed. Eight phenotypes were binary: Depression, Type 2 Diabetes (T2D), Coronary Artery Disease (CAD), Inflammatory Bowel Disease (IBD), Rheumatoid arthritis (RheuArth), Multiple Sclerosis (MultiScler), Breast Cancer, and Prostate Cancer. Three phenotypes were continuous: Intelligence, Height, and Body Mass Index (BMI). Further information regarding phenotype definitions can be found in the Supplementary Material.

Analysis was performed on a subset of ∼50,000 UKB participants for each outcome. For each continuous trait (Intelligence, Height, BMI), a random sample was selected. For disease traits, all cases were included, except for Depression and CAD where a random sample of 25,000 cases was selected. Controls were randomly selected to obtain a total sample size of 50,000. Sample sizes for each phenotype after genotype data quality control are shown in Table 1.

**Table 1.** Comparison between observed and estimated values on the absolute scale across polygenic score quantiles for binary and normally distributed phenotypes. Estimated values are based on the observed AUC/R^2^ of the polygenic score, the prevalence of binary phenotypes, and mean and standard deviation (SD) of continuous phenotypes in UKB.

| **Binary** |  |  |  |  |  |  |
| --- | --- | --- | --- | --- | --- | --- |
| **Phenotype** | **Mean Abs. Diff.** | **Mean Abs. Diff. of SD** | **N** | **Ncas** | **Ncon** | **Skewness** |
| Depression | 1.5% | NA | 49999 | 24999 | 25000 | NA |
| T2D | 2.4% | NA | 49999 | 14888 | 35111 | NA |
| CAD | 1.5% | NA | 49999 | 25000 | 24999 | NA |
| IBD | 6.8% | NA | 49999 | 3461 | 46538 | NA |
| MultiScler | 12.6% | NA | 49999 | 1137 | 48862 | NA |
| RheuArth | 6.8% | NA | 49999 | 3408 | 46591 | NA |
| Breast_Cancer | 4.6% | NA | 49999 | 8512 | 41487 | NA |
| Prostate_Cancer | 8.7% | NA | 50000 | 2927 | 47073 | NA |
| **Continuous** |  |  |  |  |  |  |
| **Phenotype** | **Mean Abs. Diff.** | **Mean Abs. Diff. of SD** | **N** | **Ncas** | **Ncon** | **Skewness** |
| Intelligence | 0.3% | 1.3% | 50000 | NA | NA | 0.144 |
| Height | 0.1% | 1.5% | 49999 | NA | NA | 0.117 |
| BMI | 0.2% | 6% | 49999 | NA | NA | 0.592 |

#### Genetic data

UKB released imputed dosage data for 488,377 individuals and ∼96 million variants, generated using IMPUTE4 software (Bycroft et al., 2018) with the Haplotype Reference Consortium reference panel (McCarthy et al., 2016) and the UK10K Consortium reference panel (UK10K Consortium, 2015). This study retained individuals that were of European ancestry based on 4-means clustering on the first 2 principal components provided by the UKB, had congruent genetic and self-reported sex, passed quality assurance tests by UKB, and removed related individuals (>3^rd^ degree relative, KING threshold > 0.044) using relatedness kinship (KING) estimates provided by the UKB (Bycroft et al., 2018). The imputed dosages were converted to hard-call format for all variants.

### Polygenic scoring

Polygenic scores were derived within a reference-standardised framework, whereby polygenic scores are derived using a common set of genetic variants, linkage disequilibrium estimates, and allele frequency estimates (Pain et al., 2020). This approach is well suited for the clinical setting and is also good practice for research purposes.

#### SNP-level QC

HapMap3 variants from the LD-score regression website (see Web Resources) were extracted from UKB, inserting any HapMap3 variants that were not available in the target sample as missing genotypes (as required for reference MAF imputation by the PLINK allelic scoring function) (Chang et al., 2015). No other SNP-level QC was performed.

#### Individual-level QC

Individuals of European ancestry were retained for polygenic score analysis. They were identified using 1000 Genomes Phase 3 projected principal components of population structure, retaining only those within three standard deviations from the mean for the top 100 principal components. This process will also remove individuals who are outliers due to technical genotyping or imputation errors.

#### GWAS summary statistics

GWAS summary statistics were identified for the phenotypes as defined above, or similar phenotypes (descriptive statistics in Table S1), excluding GWAS with documented sample overlap with the target sample of UKB. GWAS summary statistics underwent quality control to extract HapMap3 variants, remove ambiguous variants, remove variants with missing data, flip variants to match the reference retain variants with a minor allele frequency (MAF) > 0.01 in the European subset of 1KG Phase 3, retain variants with a MAF > 0.01 in the GWAS sample (if available), retain variants with a INFO > 0.6 (if available), remove variants with a discordant MAF (>0.2) between the reference and GWAS sample (if available), remove variants with association p-values >1 or </=0, remove duplicate variants, and remove variants with sample size >3SD from the median sample size (if per variant sample size is available).

#### Reference genotype datasets

Target sample genotype-based scoring was standardised using the European subset of 1000 Genomes Phase 3 (N=503).

#### Polygenic scoring methodology

Polygenic scoring was carried out using DBSLMM (Yang & Zhou, 2020), which models LD between genetic variants and applies shrinkage parameters to avoid overfitting. DBSLMM is a computationally scalable method that performs similarly to other leading polygenic scoring methods (Pain et al., 2020). For comparison with the DBSLMM polygenic score results, a threshold and clump (pT+clump) approach was also used.

pT+clump was performed using an *R*^2^ threshold of 0.1 and window of 250kb. Within the MHC region (28-34Mb on chromosome 6), the pT+clump method retains only the single most significant variant due to long range and complex LD in this region. Ten p-value thresholds were used to select variants: 1×10^−8^, 1×10^−6^, 1×10^−4^, 1×10^−2^, 0.1, 0.2, 0.3, 0.4, 0.5 and 1.

After preparation of GWAS summary statistics, polygenic scores were calculated using PLINK with reference MAF imputation of missing data. All scores were standardized (scaled and centered) based on the mean and standard deviation of polygenic scores in the reference sample.

### Converting from relative to absolute scale

Converting to the absolute scale here requires updating the distribution parameters for a phenotype, given that we observe a polygenic score within a specified range (determined by quantiles). We develop methodology to achieve this for both binary and normally distributed phenotypes, using only the distribution of the phenotype and the predictive utility of the polygenic score.

Broadly, our approach defines the population distribution for the polygenic score using a measure of its predictive utility within normal distribution theory. The polygenic score quantiles are then estimated and, using these and the phenotype and polygenic score distributions, we derived the required, updated distribution parameters for the phenotype using conditional probability rules.

For binary phenotypes, such as major depression and multiple sclerosis, polygenic scores can be modelled as a mixture of two normal distributions, using the population prevalence of the phenotype, and the predictive utility of the polygenic scores, often indicated by the area-under-the-ROC-curve (AUC). Once this distribution has been defined, the quantiles of the polygenic scores, and the proportion of cases within each quantile is estimated. Quantile estimation for the mixture distribution requires using a root-finding algorithm; here we use the ‘uniroot’ function in the ‘base’ R package (R Core Team, 2015). A full derivation of the formulae for conversion to the absolute scale is available in the Supplementary Material.

For normally distributed continuous phenotypes, such as height and IQ, polygenic scores are defined as part of a bivariate normal distribution with the phenotype, using the mean and standard deviation of the phenotype in the general population, and the predictive utility of the polygenic scores, often indicated as the variance explained (*R*^2^). Once this distribution has been defined, the quantiles of the polygenic scores, and the phenotype mean and standard deviation within each quantile is estimated. The mean and standard deviation of the phenotype within each polygenic score quantile are estimated using the ‘mtmvnorm’ function in the ‘tmvtnorm’ R package (Wilhelm & Manjunath, 2015). A full derivation of the formulae for the conversion to the absolute scale is available in the Supplementary Material.

Both conversions use some assumption of normality when defining the distribution of the polygenic score, which is underpinned by the central limit theorem. We note that polygenic scores derived using the pT+clump approach will often include only a small number of genetic variants when using a stringent p-value threshold and may therefore not fit a normal distribution. To determine whether the conversions are biased in this scenario, we compared absolute estimates to those observed when using pT+clump polygenic scores based on the most stringent p-values threshold available, whilst retaining at least 5 genetic variants.

### Estimating predictive utility of polygenic scores

Two approaches were explored to estimate the predictive utility of polygenic scores.

The *pseudovalidate* function of the ‘lassosum’ R package (Mak, Porsch, Choi, Zhou, & Sham, 2017) estimates the correlation (*R*) between the polygenic score and GWAS phenotype to identify the optimal lassosum hyper parameters (s and lambda). We have previously shown lassosum has a similar predictive utility to the DBSLMM polygenic score (Pain et al., 2020). For continuous phenotypes, the variance explained by the polygenic scores is *R*^*2*^. For binary phenotypes, the AUC is obtained from the correlation via calculation of the Cohen’s D, accounting for the GWAS sampling ratio (Aaron, Kromrey, & Ferron, 1998; Rice & Harris, 2005). The Cohen’s D is calculated from *R* as:

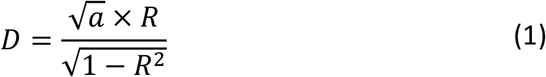

where *a* is a correction factor for imbalanced GWAS sampling ratio (*n*_1_ ≠ *n*_2_),

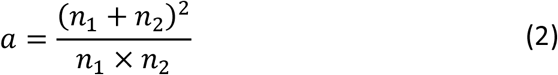

The AUC is calculated from Cohen’s D as:

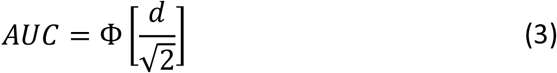

where Ф is the normal cumulative distribution function.

The ‘AVENGEME’ R package (Palla & Dudbridge, 2015) includes a function called ‘polygenescore’ which has several purposes including the estimation of the AUC/*R*^2^ of polygenic scores derived using the pT+clump approach. AVENGEME requires various input parameters, including the SNP-based heritability of the phenotype and the proportion of variants with zero effect (pi0). The SNP-based heritability was estimated from the GWAS summary statistics using LD-score regression (Bulik-Sullivan et al., 2015). However, pi0 is challenging to estimate from GWAS summary statistics, and so a range of pi0 values were used (0.92, 0.94, 0.96, 0.98).

In a third approach, we also explored the G-WIZ R package (Patron, Serra-Cayuela, Han, Li, & Wishart, 2019). This method is able to estimate the AUC of polygenic scores derived from GWAS of binary outcomes, but it cannot currently be applied to continuous outcomes. A brief investigation of the method also highlighted that substantial computational resources would be required to model polygenic scores derived using genome-wide variation, as opposed to a smaller number of genome-wide significant variants. For these reasons, we have not considered this approach further.

### Validation procedure

#### Conversion to absolute scale

Conversion to the absolute scale for binary and continuous phenotypes was validated in UKB. For binary phenotypes, the conversion was validated by comparing the observed number of affected individuals within each polygenic score quantile to the estimated values. For continuous phenotypes, the conversion was validated by comparing the observed phenotype mean and standard deviation (SD) within each polygenic score quantile to estimated values. When validating the conversion to the absolute scale, the observed AUC/R^2^ of the polygenic score, and observed sampling ratio or phenotype mean and SD were used to estimate the polygenic score distribution. We also compared observed and estimated values for height stratified by sex.

#### Estimation of polygenic score AUC/R^2^

To validate the AUC and *R*^*2*^ estimates derived from AVENGEME and lassosum, we compared the estimated values to those observed in UKB. We also used the estimated AUC/R^2^ values when converting polygenic score into absolute terms, to determine the extent to which differences between observed and estimated AUC/R^2^ values influenced the results.

### Development of interactive visualisation tool

We developed an interactive webtool for converting and visualising standardised polygenic scores to the absolute scale for binary and normally distributed phenotypes. The webtools were developed using the ‘shiny’ package in R, and are hosted on the shinyapps.io website (see URLs). For the shiny app implementation of the absolute scale conversion, the polygenic score distribution is split into 1000 quantiles to increase the precision of the results.

## Results

### Validating conversion to absolute terms

We validated the approach for converting a polygenic score into absolute terms by comparing the observed and estimated distributions of phenotypes within DBSLMM polygenic score quantiles. When the AUC/*R*^2^ of the polygenic score is known, the absolute estimates were highly concordant with observed values (Table 1, Figure 1-2).

**Figure 1.**
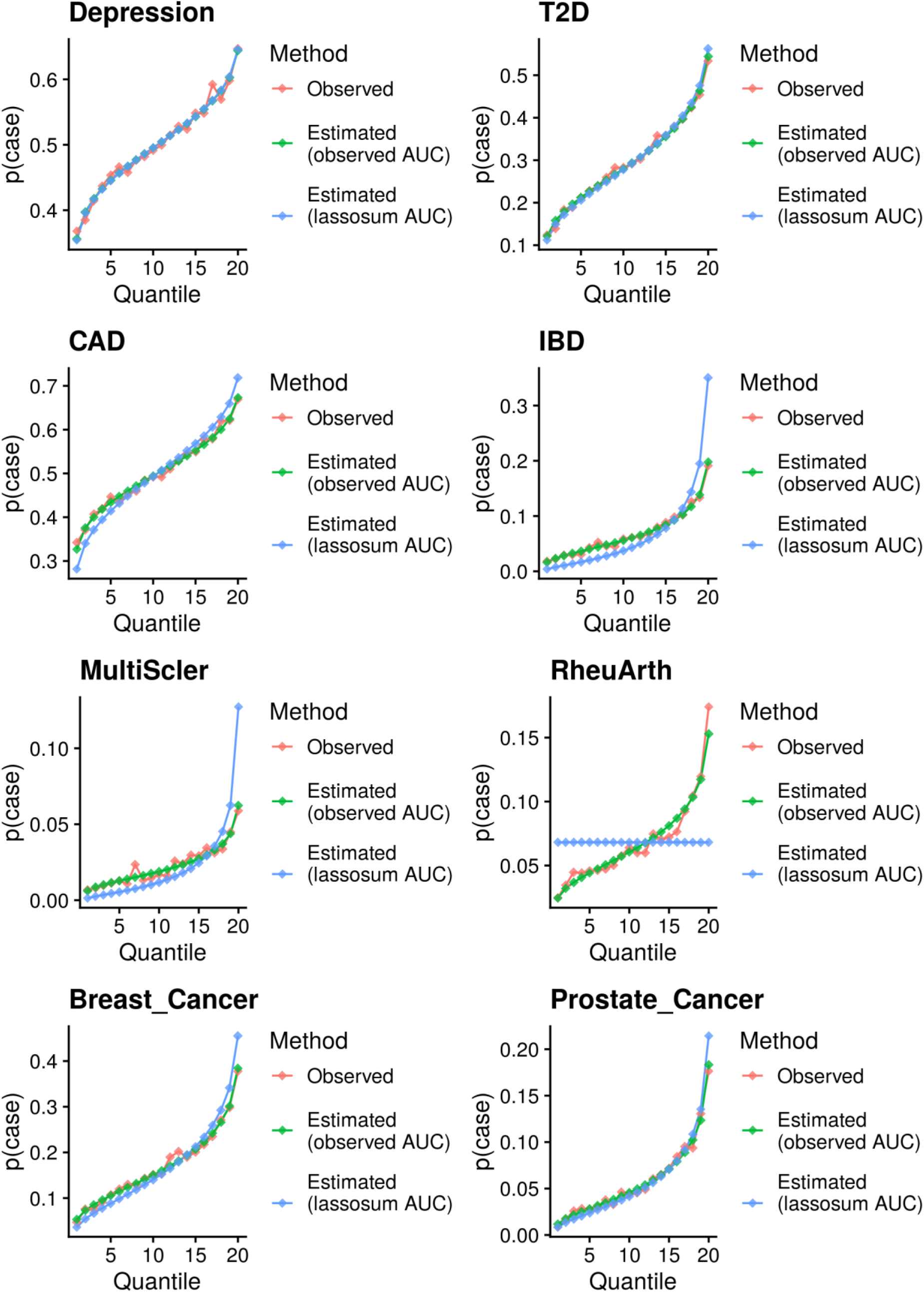
Comparison of observed and estimated probability of being a case across 20 DBSLMM polygenic score quantiles. Estimated values are based on either the observed polygenic score AUC, or the lassosum estimated AUC.

**Figure 2.**
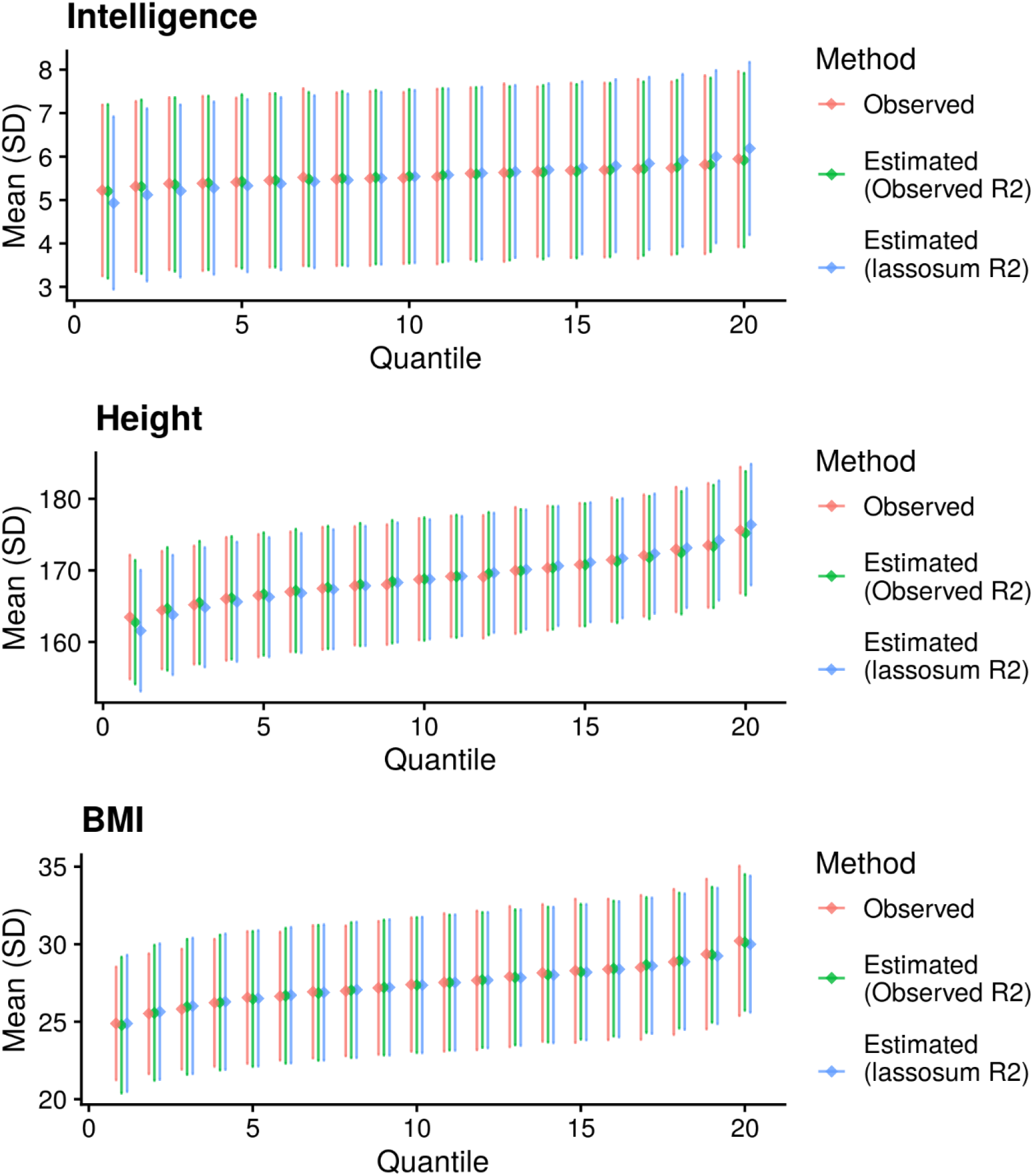
Comparison of observed and estimated phenotype mean and standard deviation across 20 DBSLMM polygenic score quantiles. Estimated values are either based on the observed polygenic score R^2^, or the lassosum estimated R^2^.

For binary phenotypes (Table 1, Figure 1), the mean absolute difference between the observed and estimated proportion of cases was between 12.6% for MultiScler and 1.5% for both Depression and CAD. The concordance between observed and estimated values decreased as the number of cases within UKB decreased, reflecting increased error in the observed values.

For the three continuous phenotypes (Table 1, Figure 2), the mean absolute difference between observed and estimated means was <0.3%. The mean absolute difference between observed and estimated phenotype standard deviations across polygenic score quantiles were 1.3% for Intelligence, 1.5% for Height, and 6% for BMI. The reduced concordance between observed and estimated values for BMI reflects the increased skewness of this phenotype in UKB.

To demonstrate the flexibility of the conversion to model absolute risk within stratified populations, we compared observed and estimated absolute values for height within males and females separately. Again, the concordance between observed and expected values was high (Figure S1). Given large sex differences in height, the polygenic score *R*^2^ increased when stratified by sex, and correspondingly the difference in mean height across polygenic score quantiles was larger, and the standard deviation of height within polygenic score quantiles was smaller.

The observed and estimated absolute values were also highly concordant when using pT+clump polygenic scores defined using stringent p-value thresholds (Figures S2-S3). Some discrepancy between observed and estimated values was present for Depression due to the low predictive utility of the polygenic score when using the stringent p-value threshold to select variants.

We have provided examples using these conversions to interpret polygenic scores for an individual (Figure 3-4). For binary phenotypes, we use the example schizophrenia, with a population prevalence of 1% and a polygenic score AUC of ∼0.67 (Pardiñas et al., 2018). If an individual has a polygenic Z-score of 1.96, they are in the 97.5^th^ percentile of schizophrenia polygenic scores. However, given the modest AUC of the polygenic score, only 2.7% of individuals with that polygenic score will develop schizophrenia (Figure 3). For continuous phenotypes, we use the example of intelligence quotient (IQ), with a population mean of 100 and standard deviation of 15, for which the educational attainment polygenic score can explain 10% of the variance (Allegrini et al., 2019). If an individual has a polygenic Z-score of −1.96, they are in the 2.5^th^ percentile of educational attainment polygenic scores. The mean IQ of individuals with this polygenic score is 90.7, with 95% prediction intervals from 62.8 to 118.6 (Figure 4).

**Figure 3.**
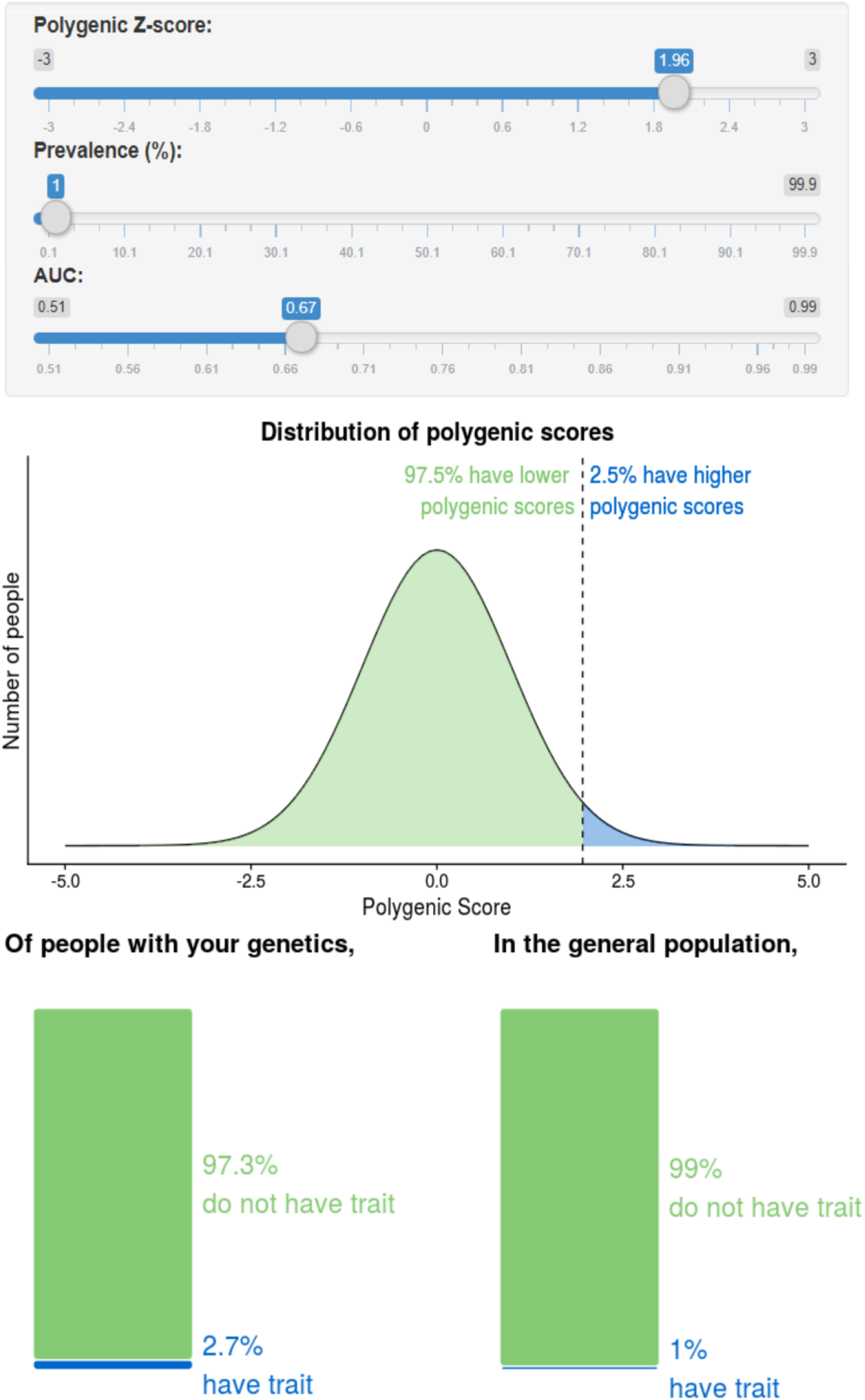
*Shiny app implementing absolute scale conversion for binary phenotypes. Parameters reflect prevalence of schizophrenia and AUC of the schizophrenia polygenic score* (Pardiñas et al., 2018).

**Figure 4.**
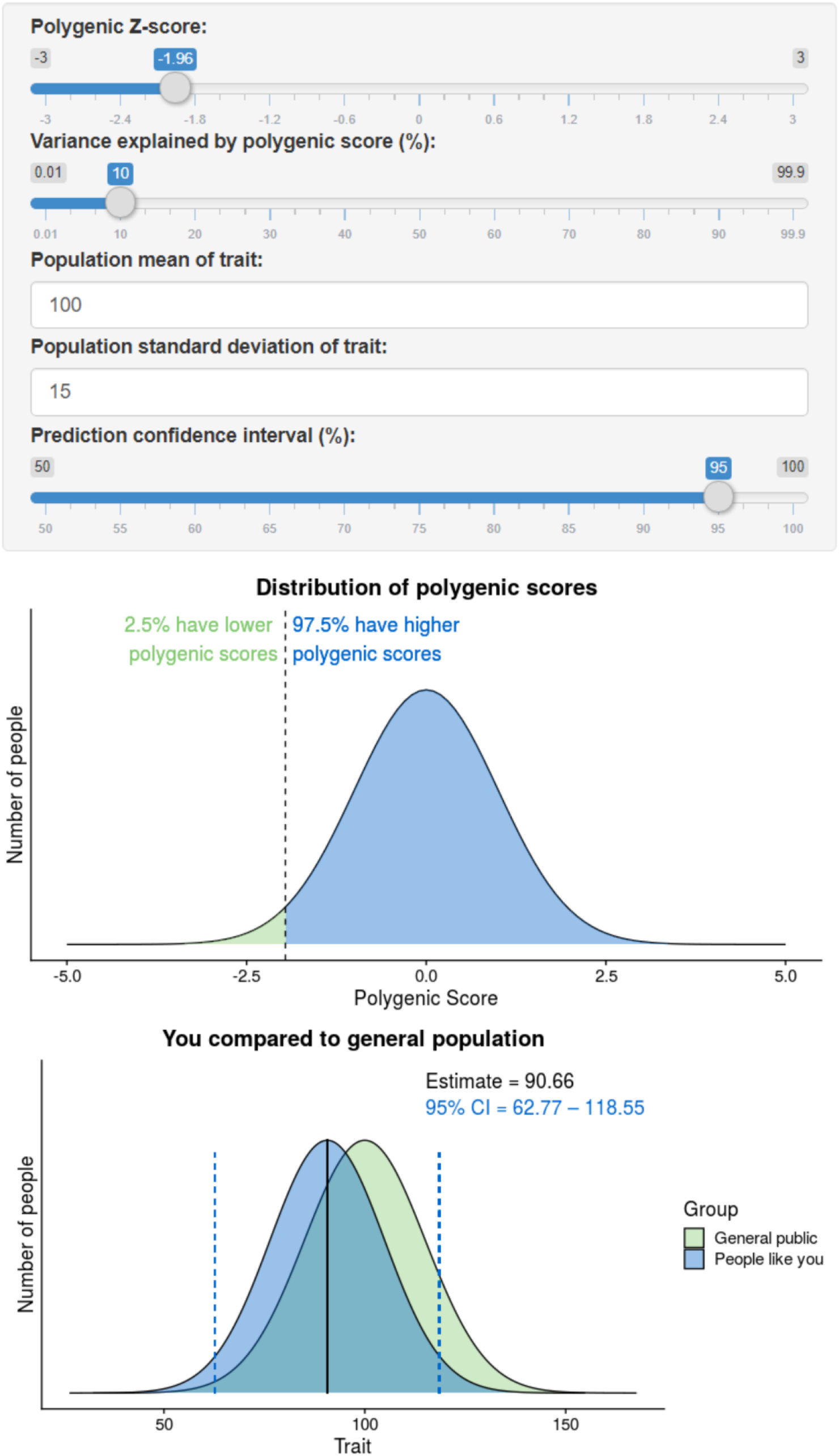
*Shiny app implementing absolute scale conversion for normally distributed phenotypes. Parameters reflect mean and SD of IQ, and R^2^ of educational attainment polygenic score for IQ* (Allegrini et al., 2019).

To illustrate the full range of predictive utility of polygenic scores on the absolute scale, we have simulated results based on a range of polygenic score AUC and prevalence values for binary phenotypes, and range of polygenic scores *R*^2^ values for continuous phenotypes (Figures S4-S5).

### Validating polygenic scores AUC/R^2^ estimation

The lassosum estimates of AUC/*R*^2^ were concordant with the observed AUC/*R*^2^ of PRScs polygenic scores for most phenotypes (Table 2). The absolute difference between estimated and observed AUC values was less than 0.04 for five of the eight binary phenotypes. The absolute difference between estimated and observed *R*^2^ values were 0.007, 0.015, and 0.046 for BMI, Intelligence and Height, respectively. The lassosum estimates were most discordant for the three autoimmune disorders included in this study. The estimated AUC was substantially higher than the observed for IBD (AUC diff = 0.115) and MultiScler (AUC diff = 0.128). The analysis did not complete for RheuArth, resulting in an AUC of 0.5 being returned by the analysis.

**Table 2.**
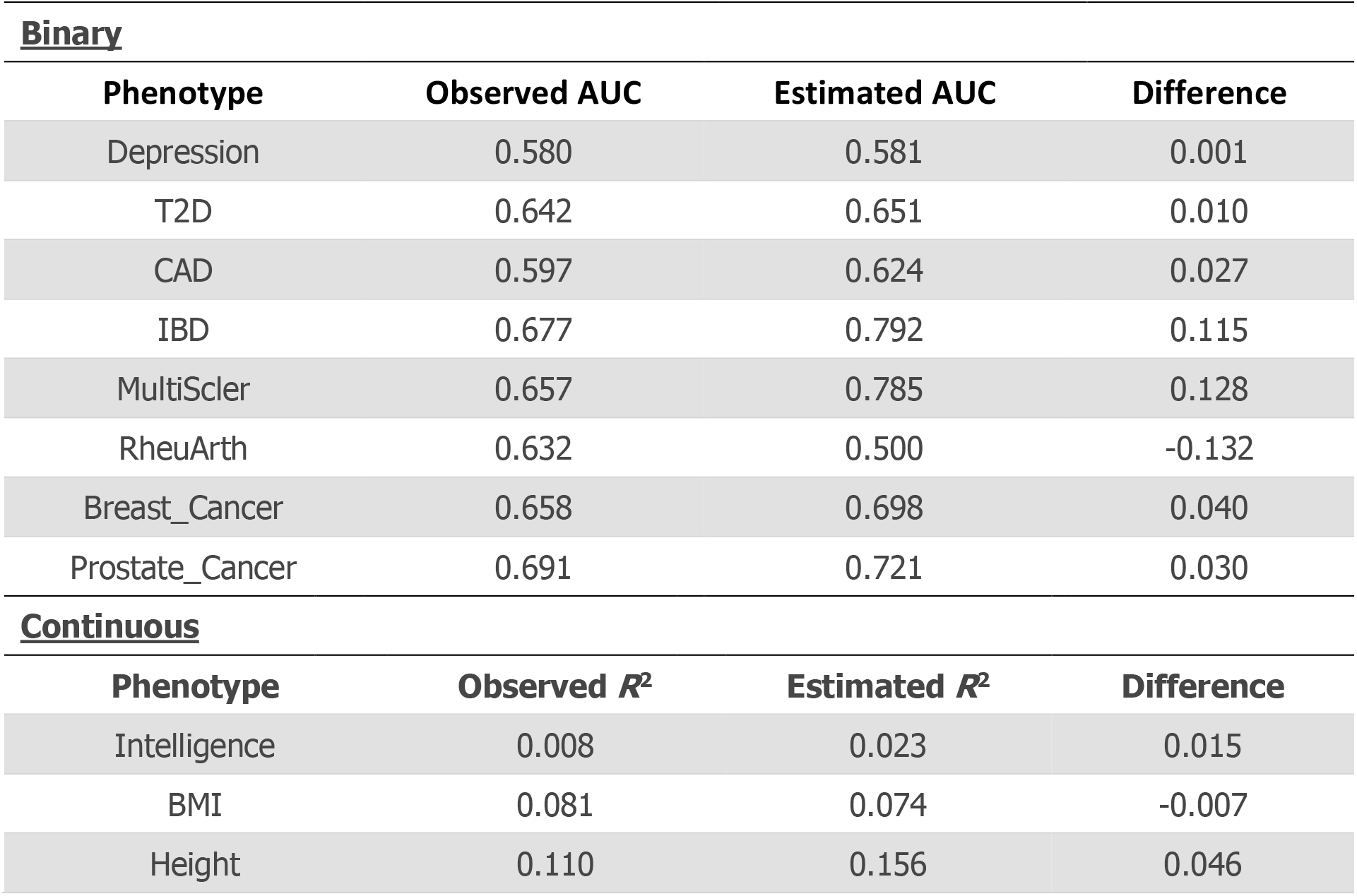
Comparison between polygenic score AUC/R^2^ observed in UKB and estimated using lassosum.

| **<u>Binary</u>** |  |  |  |
| --- | --- | --- | --- |
| **Phenotype** | **Observed AUC** | **Estimated AUC** | **Difference** |
| Depression | 0.580 | 0.581 | 0.001 |
| T2D | 0.642 | 0.651 | 0.010 |
| CAD | 0.597 | 0.624 | 0.027 |
| IBD | 0.677 | 0.792 | 0.115 |
| MultiScler | 0.657 | 0.785 | 0.128 |
| RheuArth | 0.632 | 0.500 | -0.132 |
| Breast_Cancer | 0.658 | 0.698 | 0.040 |
| Prostate_Cancer | 0.691 | 0.721 | 0.030 |
| **<u>Continuous</u>** |  |  |  |
| **Phenotype** | **Observed $R^2$** | **Estimated $R^2$** | **Difference** |
| Intelligence | 0.008 | 0.023 | 0.015 |
| BMI | 0.081 | 0.074 | -0.007 |
| Height | 0.110 | 0.156 | 0.046 |

To determine the extent to which using estimated AUC/*R*^2^ influences the absolute estimates, we compared absolute estimates derived using lassosum estimated AUC/*R*^2^, to the observed absolute values (Table 3). The results for RheuArth were highly discordant due to the incomplete analysis estimating the AUC. After excluding RheuArth, the concordance between observed and estimated absolute values remained high when using estimated AUC/*R*^2^ values. For IBD and MultiScler, the phenotypes with discordant estimates of polygenic score AUC, the mean absolute percentage difference between observed-estimated proportion of cases was 38.6% and 43.7%, respectively. Discrepancies were particularly pronounced in the upper tail of the polygenic score distribution. The ratio between estimated and observed proportions of cases in the top polygenic score quantile (top 5%) was 1.83 and 2.16 for IBD and MultiScler respectively.

**Table 3.**
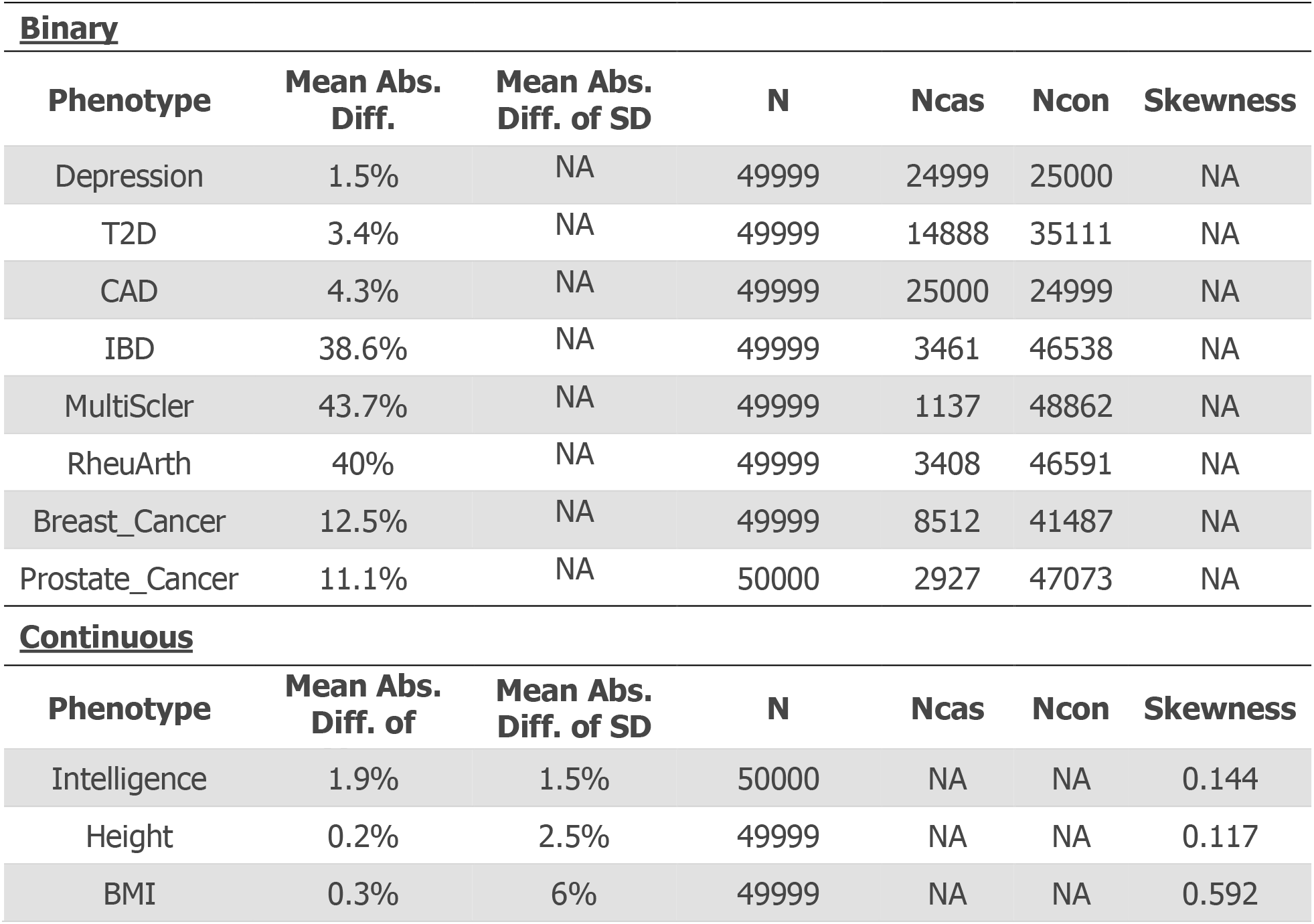
Comparison between observed and estimated case probabilities across polygenic score quantiles for binary phenotypes. Estimated values are based on the lassosum estimated AUC/R^2^ of the polygenic score.

Estimates of polygenic score AUC/*R*^2^ derived using LDSC and AVENGEME were on average less accurate than those of lassosum. Sensitivity analysis highlighted two limitations of the approach. First, that LDSC SNP-based heritability estimates were often discordant to the SNP-based heritability estimated by AVENGEME based on observed polygenic score associations. For example, LDSC estimated the multiple sclerosis SNP-based heritability to be 2% whereas the AVENGEME SNP-based heritability estimate based on observed polygenic score associations was 13.3%. This discordance between the LDSC SNP-based heritability estimate and the SNP-based heritability reflected by the polygenic score associations leads to inaccurate estimates of polygenic score AUC/*R*^2^ by AVENGEME. The second limitation is that the polygenicity of GWAS phenotypes was not known, so we assumed an average degree of polygenicity for all GWAS phenotypes. This average polygenicity assumption contributed to inaccurate AUC/*R*^2^ estimates. Results of the LDSC/AVENGEME analysis are further described in the Supplementary Material (Figure S6-S7, Table S2-S3).

## Discussion

This study has derived and evaluated methods for converting polygenic scores for both binary and normally distributed phenotypes from a relative value, of where the polygenic score lies on the distribution, into a value on the absolute scale. For a disorder, the conversion provides an estimate of the proportion of cases within a polygenic score quantile, and for a normally distributed trait, the conversion gives the trait mean and standard deviation within a polygenic score quantile. Comparison of absolute estimates with observed values within UKB show the method is highly accurate when the AUC (for disorders) or *R*^2^ (for a trait) of the polygenic score is known. Furthermore, we show that lassosum pseudovalidate function can provide accurate estimates of AUC/*R*^2^ in most instances, though inaccuracies in AUC/*R*^2^ estimation can substantially bias absolute estimates in the extremes of the polygenic score distribution.

Interpretation of polygenic scores is one of the greatest barriers to their safe application to the clinical setting, and also poses a problem when polygenic scores are delivered to the individuals via direct-to-consumer genetics testing companies. Our study provides a summary statistic-based approach for converting polygenic scores into absolute terms, enabling their accurate interpretation. Furthermore, where the predictive utility of polygenic scores is unknown, we show that lassosum pseudovalidate may be used although the results should be interpreted with caution.

The predictive utility of polygenic scores is under a great deal of scrutiny. A major criticism is that they are only predictive at the group-level, i.e. for research, but are poor predictors at the individual-level. Our approach of converting polygenic scores to the absolute scale helps understand this distinction as it converts the group-level metric of predictive utility (AUC or *R*^2^) into an absolute value for an individual (risk of disorder or trait value with prediction intervals). Indeed, the absolute estimates do not vary substantially across the polygenic score distribution except at the extremes, which reflects the modest AUC/*R*^2^ of the current polygenic scores. The predictive utility of polygenic scores will increase with more powerful GWAS and better resolution of the causal variant. However, these scores will always be probabilistic, and will never give deterministic predictions as genetic variation only explains part of the phenotypic variance. Polygenic scores can be combined with other risk factors to increase the accuracy of prediction, though the value of prediction also depends partly on the actions or interventions available to address the predicted outcome.

We have developed an interactive webtool implementing these methods to convert polygenic scores into absolute risk or trait estimates, also providing visual aids to promote the accurate interpretation of differences in risk. We also plan to implement these methods on Impute.me, a popular non-profit website providing polygenic scores for users who upload genotype data from a direct-to-consumer genetic testing company.

There are several limitations of this study. Firstly, this summary statistic-based approach relies on the input parameters being representative of the target individual’s demographic. This study focuses on UKB participants of European ancestry, and we define the distribution of the phenotype using the observed distribution within UKB. However, specifying the distribution of a phenotype and the AUC/*R*^2^ of the polygenic score for a population that is representative of the target individual may be challenging. For example, assuming the phenotype distribution and polygenic score AUC/R^2^ found in a European population will give biased results in non-European populations where differences exist in the phenotype distribution and polygenic score AUC/*R*^2^ (T. R. Wood & Owens, 2020). Most GWAS are performed in European populations, and the variance explained by polygenic scores is typically higher in Europeans than non-Europeans (Martin et al., 2019). Secondly, although the lassosum pseudovalidate approach works well in most instances, it can provide inaccurate results which could lead to inaccurate absolute estimates. Further development of methods that can estimates the AUC/*R*^2^ of polygenic scores without a validation sample would be useful. Alternatively, the AUC/R^2^ could be based upon resources such as The Polygenic Score (PGS) Catalog (Lambert et al., 2021), which collate polygenic score data and corresponding prediction metrics. To support this effort, future polygenic score risk prediction studies should follow suggested reporting standards (Wand et al., 2021). Thirdly, for continuous phenotypes, the approach is tailored to normally distributed phenotypes. Further development of methods to account for non-normally distributed phenotypes may be useful. Finally, for binary traits or disease, we calculate life-time risks that are based on pre-specified prevalence, which do not account for the risk period an individual has already lived through or other risk factors.

In summary, this study has provided an approach for converting polygenic scores into absolute risk and predictions based on GWAS summary statistics. It establishes a framework for appropriate and accurate interpretation of polygenic scores by patients, consumers, and healthcare professionals.

## Data Availability

Individual-level data for UK Biobank must be applied for via the Access Management System (https://www.ukbiobank.ac.uk/enable-your-research/apply-for-access). All code used in this study is publicly available (https://opain.github.io/GenoPred/).

## URLS

- Interactive webtool/shiny apps: https://opain.github.io/GenoPred/PRS_to_Abs_tool.html
- LDSC HapMap 3 SNP-list: https://data.broadinstitute.org/alkesgroup/LDSCORE/w_hm3.snplist.bz2
- Impute.me: https://impute.me/
- GenoPred website: https://opain.github.io/GenoPred

## Acknowledgements

This paper represents independent research funded by the UK Medical Research Council (MR/N015746/1), and the National Institute for Health Research (NIHR) Biomedical Research Centre at South London and Maudsley NHS Foundation Trust and King’s College London. The authors acknowledge use of the research computing facility at King’s College London, Rosalind (https://rosalind.kcl.ac.uk), which is delivered in partnership with the NIHR Maudsley BRC, and part-funded by capital equipment grants from the Maudsley Charity (award 980) and Guy’s & St. Thomas’ Charity (TR130505). The views expressed are those of the authors and not necessarily those of the NHS, the NIHR or the Department of Health and Social Care.

UKB: This research was conducted under UK Biobank application 18177.

## References

Aaron, B., Kromrey, J. D., & Ferron, J. (1998). Equating” r”-based and” d”-based effect size indices: problems with a commonly recommended formula. ERIC Clearinghouse.

Allegrini, A. G., Selzam, S., Rimfeld, K., von Stumm, S., Pingault, J.-B., & Plomin, R. (2019). Genomic prediction of cognitive traits in childhood and adolescence. Molecular Psychiatry, 24(6), 819–827.

Bulik-Sullivan, B. K., Loh, P.-R., Finucane, H. K., Ripke, S., Yang, J., Patterson, N.,… Consortium, S. W. G. of the P. G. (2015). LD Score regression distinguishes confounding from polygenicity in genome-wide association studies. Nature Genetics, 47(3), 291–295.

Bycroft, C., Freeman, C., Petkova, D., Band, G., Elliott, L. T., Sharp, K.,… O’Connell, J. (2018). The UK Biobank resource with deep phenotyping and genomic data. Nature, 562(7726), 203–209.

Chang, C. C., Chow, C. C., Tellier, L. C. A. M., Vattikuti, S., Purcell, S. M., & Lee, J. J. (2015). Second-generation PLINK: rising to the challenge of larger and richer datasets. Gigascience, 4(1), 1.

Choi, S. W., Mak, T. S.-H., & O’Reilly, P. F. (2020). Tutorial: a guide to performing polygenic risk score analyses. Nature Protocols, 1–14.

Furlotte, N. A., Kleinman, A., Smith, R., & Hinds, D. (2015). White paper 23-12: Estimating Complex Phenotype Prevelance Using Predictive Models.

Gigerenzer, G., Gaissmaier, W., Kurz-Milcke, E., Schwartz, L. M., & Woloshin, S. (2007). Helping doctors and patients make sense of health statistics. Psychological Science in the Public Interest, 8(2), 53–96.

Howard, D. M., Adams, M. J., Clarke, T.-K., Hafferty, J. D., Gibson, J., Shirali, M.,… Wigmore, E. M. (2019). Genome-wide meta-analysis of depression identifies 102 independent variants and highlights the importance of the prefrontal brain regions. Nature Neuroscience, 22(3), 343–352.

Khera, A. V, Chaffin, M., Aragam, K. G., Haas, M. E., Roselli, C., Choi, S. H.,… Ellinor, P. T. (2018). Genome-wide polygenic scores for common diseases identify individuals with risk equivalent to monogenic mutations. Nature Genetics, 50(9), 1219–1224.

Lambert, S. A., Gil, L., Jupp, S., Ritchie, S. C., Xu, Y., Buniello, A.,… Parkinson, H. (2021). The Polygenic Score Catalog as an open database for reproducibility and systematic evaluation. Nature Genetics, 1–6.

Lewis, C. M., & Vassos, E. (2020). Polygenic risk scores: from research tools to clinical instruments. Genome Medicine, 12, 1–11.

Mak, T. S. H., Porsch, R. M., Choi, S. W., Zhou, X., & Sham, P. C. (2017). Polygenic scores via penalized regression on summary statistics. Genetic Epidemiology, 41(6), 469–480.

Martin, A. R., Kanai, M., Kamatani, Y., Okada, Y., Neale, B. M., & Daly, M. J. (2019). Clinical use of current polygenic risk scores may exacerbate health disparities. Nature Genetics, 51(4), 584–591.

McCarthy, S., Das, S., Kretzschmar, W., Delaneau, O., Wood, A. R., Teumer, A.,… Sharp, K. (2016). A reference panel of 64,976 haplotypes for genotype imputation. Nature Genetics.

Pain, O., Glanville, K. P., Hagenaars, S. P., Selzam, S. P., Fürtjes, A. E., Gaspar, H. A.,… Lewis, C. M. (2020). Evaluation of Polygenic Prediction Methodology within a Reference-Standardized Framework. BioRxiv.

Palla, L., & Dudbridge, F. (2015). A fast method that uses polygenic scores to estimate the variance explained by genome-wide marker panels and the proportion of variants affecting a trait. The American Journal of Human Genetics, 97(2), 250–259.

Pardiñas, A. F., Holmans, P., Pocklington, A. J., Escott-Price, V., Ripke, S., Carrera, N.,… Hamshere, M. L. (2018). Common schizophrenia alleles are enriched in mutation-intolerant genes and in regions under strong background selection. Nature Genetics, 50(3), 381.

Patron, J., Serra-Cayuela, A., Han, B., Li, C., & Wishart, D. S. (2019). Assessing the performance of genome-wide association studies for predicting disease risk. PloS One, 14(12), e0220215.

Polderman, T. J. C., Benyamin, B., de Leeuw, C. A., Sullivan, P. F., van Bochoven, A., Visscher, P. M., & Posthuma, D. (2015). Meta-analysis of the heritability of human traits based on fifty years of twin studies. Nature Genetics, 47(7), 702–709. https://doi.org/10.1038/ng.3285

R Core Team. (2015). R: A Language and Environment for Statistical Computing. Vienna, Austria. Retrieved from http://www.r-project.org

Rice, M. E., & Harris, G. T. (2005). Comparing effect sizes in follow-up studies: ROC Area, Cohen’s d, and r. Law and Human Behavior, 29(5), 615.

UK10K Consortium. (2015). The UK10K project identifies rare variants in health and disease. Nature, 526(7571), 82–90.

Wand, H., Lambert, S. A., Tamburro, C., Iacocca, M. A., O’Sullivan, J. W., Sillari, C.,… Brockman, D. (2021). Improving reporting standards for polygenic scores in risk prediction studies. Nature, 591(7849), 211–219.

Wilhelm, S., & Manjunath, G. B. (2015). tmvtnorm: Truncated Multivariate Normal and Student t Distribution.

Wood, A. R., Esko, T., Yang, J., Vedantam, S., Pers, T. H., Gustafsson, S.,… Kutalik, Z. (2014). Defining the role of common variation in the genomic and biological architecture of adult human height. Nature Genetics, 46(11), 1173.

Wood, T. R., & Owens, N. (2020). Using synthetic datasets to bridge the gap between the promise and reality of basing health-related decisions on common single nucleotide polymorphisms. F1000Research, 8(2147), 2147.

Wray, N. R., Lin, T., Austin, J., McGrath, J. J., Hickie, I. B., Murray, G. K., & Visscher, P. M. (2021). From basic science to clinical application of polygenic risk scores: a primer. JAMA Psychiatry, 78(1), 101–109.

Yang, S., & Zhou, X. (2020). Accurate and scalable construction of polygenic scores in large biobank data sets. The American Journal of Human Genetics, 106(5), 679–693.

Zipkin, D. A., Umscheid, C. A., Keating, N. L., Allen, E., Aung, K., Beyth, R.,… Korenstein, D. (2014). Evidence-based risk communication: a systematic review. Annals of Internal Medicine, 161(4), 270–280.

