## Supplementary Text and Figures for "A Tool for Translating Polygenic Scores onto the Absolute Scale Using Summary Statistics"

### Supplementary material for ‘**Improving interpretability of polygenic scores using only summary statistics’ by Pain et al.**

#### UKB Outcome definitions

*Depression.* UKB participants were coded as depression cases if they met the Composite International Diagnostic Interview Short Form criteria for lifetime depression which was assessed in the online Mental Health Questionnaire (MHQ) using scoring protocols proposed by Davis et al (Davis et al., 2020). Depression cases were screened for indications of schizophrenia or bipolar disorder according to the MHQ. Controls excluded if they show any psychiatric indications according to the MHQ or depression indications according to: ICD-10 diagnoses; endorsement of self-reported depression; endorsement of current antidepressant usage; single or current depression according to the criteria adopted by Smith, et al (Smith et al., 2013). Further details of the exclusion criteria have been previously described (Glanville et al., 2020).

*T2D.* Cases were identified based on a combination of hospital episode statistics, using both ICD-9 and ICD-10, the national death register, and self-reported questionnaire data. In order to classify as a case for type 2 diabetes, self-reported type 2 or generic diabetes status was established in the nurse interview and the touchscreen questionnaire. However, participants were only classified as cases when they reported in the questionnaire that they had not been treated with insulin in the first year after diagnosis and had been diagnosed after the age of 35 years. Type 2 diabetes controls did not fulfil these criteria and did not have any other types of diabetes. Further details of the T2D definition have been previously published (Fürtjes, Coleman, Tyrrell, Lewis, & Hagenaars, 2020).

*Coronary artery disease (CAD).* Participants who were registered in the hospital in-patient data or the death register to have had ischemic heart diseases, or participants who had coronary revascularization operations were classified as coronary artery disease cases in this study. If participants self-reported those conditions in the nurse interview or the touchscreen questionnaire, they were also considered to have coronary artery disease. Coronary artery disease controls did not fulfil those criteria. Further details of the CAD definition have been previously published (Fürtjes et al., 2020).

*Autoimmune diseases (IBD, RheuArth, MultiScler):* UKB participants were coded as autoimmune cases if at least two of the following measures were observed: ICD-10 diagnoses from Hospital Episode Statistics; endorsement of self-reported autoimmune diseases; endorsement of prescription medication for the corresponding autoimmune diseases. More than one hospital admission for the respective autoimmune conditions was also sufficient. Controls were excluded if any of the following were observed: Pernicious Anemia, Autoimmune Thyroid Disease, Type 1 diabetes, Multiple Sclerosis, Myasthenia Gravis, Coeliac, Inflammatory Bowel Disease, Hidradenitis Suppurativa, Pemphigoid/Pemphigus, Psoriasis, Ankylosing Spondylitis, Polymyalgia Rheumatica/Giant Cell Arteritis, Psoriatic Arthritis, Rheumatoid Arthritis, Sjögren Syndrome, Systemic Lupus Erythematosus.

*Intelligence* was defined using the Fluid intelligence score variable. Fluid intelligence was assessed using the 13 item UKB Touch-screen Fluid intelligence test (Sudlow et al., 2015). The test measures the capacity to solve problems that require logic and reasoning ability, independent of acquired knowledge. The fluid intelligence variable was derived by UKB as an unweighted sum of the number of correct answers, assigning a score of 0 to unanswered questions.

*Height* was defined using the Standing height variable (Field ID: f.50.0.0).

*BMI* was defined using the Body mass index variable (Field ID: f.21001.0.0).

Breast Cancer and Prostate Cancer were defined using the self-reported illness codes (1044 = prostate cancer, 1002 = breast cancer, Field ID: f.20001).

#### Converting relative estimates into absolute terms

##### Binary traits

**Aim:** We wish to estimate the risk of disease for an individual given their polygenic score falls within a certain group of the population distribution, defined by quantiles, using only summary statistic data; specifically, the disease prevalence in the population understudy and the AUC (area under the ROC curve, here measuring how useful the polygenic score is at classifying individuals as cases or controls).

We achieve this aim in three steps. Firstly, we use the summary statistics to define the distribution of the polygenic score in the case subpopulation, the control subpopulation and the overall population. Secondly, we use the distribution of the polygenic score in the population to define the quantiles (the cut points defining the groups). Thirdly, we use the polygenic score distribution in the case (control) subpopulation to derive the required probability (the probability of being a case or control given the polygenic score falls within a certain group).

**Step 1: Defining the conditional and unconditional polygenic score distributions**

Let $Y_{i}$ denote the random disease outcome variable for individual $i$ such that $Y_{i}=1$ if the individual is a case and $Y_{i}=0$ if the individual is a control. The distribution of $Y_{i}$ is defined as:

$$Y_{i}\sim Binom\left( 1, K \right)$$

where $K=p\left( Y_{i}=1 \right)$ is the disease prevalence.

Let $X_{i}$ denote the random polygenic score variable for individual $i$. If individual $i$ is a control, then we assume that their polygenic score follows a standard normal distribution:

$$X_{i}|\left\{ Y_{i}=0 \right\}\sim N\left( 0, 1 \right)$$

If individual $i$ is a case, then we assume that their polygenic score is also normally distributed but with a different mean:

$$X_{i}|\left\{ Y_{i}=1 \right\}\sim N\left( d, 1 \right)$$

where $d$ is Cohen’s D, which measures the standardised difference between two means (here it is the standardised difference between the expected polygenic score for cases and controls).

Then, the distribution for $X_{i}$ is a *mixture* of the above two normal distributions, weighted by the disease prevalence, with the probability density function (PDF) for $X_{i}$ defined as:

$$f_{X_{i}}\left( x_{i} \right)=Kf_{X_{i}|Y_{i}}\left( x_{i}|1 \right)+(1-K)f_{X_{i}|Y_{i}}\left( x_{i}|0 \right)$$

where $f_{X_{i}|Y_{i}}\left( x_{i1}|1 \right)=\frac{1}{\sqrt{2\pi}}e^{-\frac{1}{2}{(x_{i}-d)}^{2}}$ is the PDF for the conditional polygenic score random variable given the individual is case ($X_{i}|\left\{ Y_{i}=1 \right\}$).

Similarly, $f_{X_{i}|Y_{i}}\left( x_{i}|0 \right)=\frac{1}{\sqrt{2\pi}}e^{-\frac{1}{2}{x_{i}}^{2}}$ is the PDF for the conditional polygenic score random variable given the individual is a control ($X_{i}|\left\{ Y_{i}=0 \right\}$).

Rice and Harris (Rice & Harris, 2005) showed that Cohen’s D can be approximated using:

$$d\approx\sqrt{2}\Phi^{-1}[AUC]$$

Therefore, the above distributions for the polygenic score can be defined using the AUC and the disease prevalence.

**Step 2: Defining the quantiles**

To define the required polygenic score groups we need to calculate the quantiles; the $(n-1)$ cut points that split the data into $n$ equally sized groups. The quantiles are defined in the overall population encompassing both cases and controls and we therefore need to use to unconditional polygenic score distribution (which is a mixture of two normal distributions).

Let:

- $p_{q}=q/n$ be the probability value defining a quantile boundary, and,
- $t_{q}$ be the polygenic score value corresponding to $p_{q}$;

such that:

$$p_{q}=p\left( X_{i}<t_{q} \right)$$

for $q=1,\cdots,n-1$.

Since $X_{i}$ is a mixture of two normal distributions, this becomes:

$$p_{q}=Kp\left( X_{i} | \left\{ Y_{i}=1 \right\}< t_{q} \right)+\left( 1-K \right)p\left( X_{i} | \left\{ Y_{i}=0 \right\}< t_{q} \right)$$

$$=Kp\left( Z<t_{q}-d \right)+\left( 1-K \right)p\left( Z<t_{q} \right)$$

$$=K\Phi\left[ t_{q}-d \right]+\left( 1-K \right)\Phi[t_{q}]$$

Variables $p_{q}$, $K$ and $d$ are known. The only unknown in the above equation is $t_{q}$, which can be found by solving this equation using numerical methods. For example, we used the uniroot function within R, which finds the root (solution) of an equation by searching a specified interval for the parameter that is required to be calculated and outputting the value that gives a 0 solution to the equation. The user therefore needs to input the equation such that it equals 0. In this case, that is:

$$K\Phi\left[ t_{q}-d \right]+\left( 1-K \right)\Phi\left[ t_{q} \right]-p_{q}=0$$

This will need to be repeated to calculate all quantiles.

**Step 3: Equations for the required case and control probabilities**

We now wish to derive the probability that individual $i$ is a case or a control given they have a polygenic score within a certain range (defined by the quantiles). Starting with the probability of being a case, we wish to calculate:

$$p(Y_{i}=1|t_{q-1}<X_{i}<t_{q})$$

for $q=1,\ldots,21$, where: 1. $t_{0}=-\infty$, 2. $t_{21}=\infty$, and 3. the remaining values of $t_{q-1}$ and $t_{q}$ are calculated using step 2.

Using rules of conditional probability we write:

$$p\left( Y_{i}=1 | t_{q-1}<X_{i}<t_{q} \right)=\frac{p\left( t_{q-1}<X_{i}<t_{q} | Y_{i}=1 \right)p\left( Y_{i}=1 \right)}{p\left( t_{q-1}<X_{i}<t_{q} \right)}$$

$t_{q-1}$ and $t_{q}$ define consecutive quantiles. Therefore $p\left( t_{q-1}<X_{i}<t_{q} \right)=1/n$ is a constant value for all $q=1, \ldots, (n+1)$, where $n$ is the number of groups the quantiles define. Additionally, $p\left( Y_{i}=1 \right)=K$. Therefore:

$$p\left( Y_{i}=1 | t_{q-1}<X_{i}<t_{q} \right)=nK p\left( t_{q-1}<X_{i}<t_{q} | Y_{i}=1 \right)$$

Recall that $E\left[ X_{i} | Y_{i}=1 \right]=d$, and so:

$$p\left( Y_{i}=1 | t_{q-1}<X_{i}<t_{q} \right)=nK\left( p\left( Z<t_{q}-d \right)-p\left( Z<t_{q-1}-d \right) \right)$$

$$=nK\left( \Phi\left[ t_{q}-d \right]-\Phi\left[ t_{q-1}-d \right] \right)$$

**Eq 1**

where $Z\sim N(0,1)$ and:

$$\Phi\left[ x \right]=\int_{-\infty}^{x} \frac{1}{\sqrt{2\pi}}e^{-\frac{1}{2}s^{2}}ds$$

is the cumulative distribution function (CDF) for the standard normal distribution.

The probability of being a control given the polygenic score sits within a certain range (defined by the quantiles) is derived in a similar way giving:

$$p\left( Y_{i}=0 | t_{q-1}<X_{i}<t_{q} \right)=nK\left( \Phi\left[ t_{q} \right]-\Phi\left[ t_{q-1} \right] \right)$$

**Eq 2**

The probabilities given in *Eq 1* and *Eq 2* can be used absolute risks, or to calculate further summary measures such as quantile relative risks or odds ratios.

##### Normally distributed traits

**Aim:** To estimate the mean (and variance) of the outcome trait for individuals with polygenic score within a group, defined by quantiles, using only summary statistics. The summary statistic used here is the variance in outcome explained by the polygenic score, $R^{2}$.

To achieve this aim we need to: 1. define the joint distribution of the outcome trait and the polygenic score, 2. calculate the quantiles, and 3. use this joint distribution and the quantiles to estimate the required distribution parameters for outcome conditional on the polygenic score belonging to a given group.

**Step 1: Defining the joint distribution of the outcome trait and the polygenic score**

For simplicity, let us assume that the outcome trait, $Y_{i}$, and the polygenic score, $X_{i}$, are standardised and follow a bivariate normal distribution defined as:

$$\left[ \begin{matrix} Y_{i} \\ X_{i} \end{matrix} \right]\sim N\left( \underline{\mu}, \Sigma\right)$$

where $\underline{\mu}=\left[ \begin{matrix} 0 \\ 0 \end{matrix} \right], \Sigma=\left[ \begin{matrix} 1 & R \\ R & 1 \end{matrix} \right]$ and $R^{2}$ is the variance in outcome explained by the polygenic score.

**Step 2: Defining the quantiles**

Recall:

$$p_{q}=p\left( X_{i}<t_{q} \right)=\frac{q}{n}$$

where:

- $p_{q}$ is the probability value defining a quantile (cut point) such that the data is split into $n$ equally sized groups, and,
- $t_{q}$ is the polygenic score value corresponding to $p_{q}$.

Here, polygenic score quantile boundaries are calculated using the univariate polygenic score distribution: $X_{i}\sim N\left( 0,1 \right)$. Therefore:

$$p_{q}=\Phi\left[ t_{q} \right]$$

and:

$$t_{q}=\Phi^{-1}\left[ p_{q} \right]$$

for $q=1,\ldots, n-1$.

**Step 3: Estimating the expected value of the outcome trait given the polygenic score falls within a quantile**

The principal value of interest is the expected value of the outcome trait given the polygenic score belongs to a certain quantile; that is, $E[Y_{i}|{\{t}_{q-1}<X_{i}<t_{q}\}]$.

To find this we need to solve the following:

$$E[Y_{i}|{\{t}_{q-1}<X_{i}<t_{q}\}]=\int_{-\infty}^{+\infty} y\int_{t_{q-1}}^{t_{q}} f_{Y_{i},X_{i}}\left( y,x \right)dxdy$$

where:

$$f_{Y_{i},X_{i}}\left( y,x \right)=\frac{1}{\sqrt{\left( 2\pi\right)^{2}\left| \Sigma\right|}}exp\left( -\frac{1}{2}\left[ \begin{matrix} y & x \end{matrix} \right]\Sigma^{-1}\left[ \begin{matrix} y \\ x \end{matrix} \right] \right)$$

$$=\frac{1}{2\pi\sqrt{1-R^{2}}}exp\left( -\frac{1}{2(1-R^{2})}\left( y^{2}-2Rxy+x^{2} \right) \right)$$

is the joint (bivariate normal) probability distribution function for the outcome trait random variable ($Y_{i}$) and the polygenic score random variable ($X_{i}$). $\left| \Sigma\right|$ is the determinant of the covariance matrix $\Sigma$.

Solving such an integral is done using numerical methods. Here we use the ‘mtmvnorm’ function in the ‘tmvtnorm’ R package (Wilhelm & Manjunath, 2015), which outputs:

$$\underline{\mu}^{'}=\left[ \begin{matrix} E[Y_{i}|{\{t}_{q-1}<X_{i}<t_{q}\}] \\ E[X_{i}|{\{t}_{q-1}<X_{i}<t_{q}\}] \end{matrix} \right]$$

and

$$\Sigma^{'}=\left[ \begin{matrix} Var[Y_{i}|{\{t}_{q-1}<X_{i}<t_{q}\}] & Cov\left[ Y_{i} | {\{t}_{q-1}<X_{i}<t_{q} \right\},X_{i}|{\{t}_{q-1}<X_{i}<t_{q}\}] \\ Cov\left[ Y_{i} | {\{t}_{q-1}<X_{i}<t_{q} \right\},X_{i}|{\{t}_{q-1}<X_{i}<t_{q}\}] & Var[X_{i}|{\{t}_{q-1}<X_{i}<t_{q}\}] \end{matrix} \right]$$

#### Estimation of polygenic score AUC/R2 using AVENGEME/LDSC

The ‘AVENGEME’ R package (Palla & Dudbridge, 2015) includes a function called ‘polygenescore’ which has several purposes including the estimation of the AUC/*R*^2^ of polygenic scores derived using the pT+clump approach. AVENGEME requires various input parameters, including the SNP-based heritability of the phenotype and the proportion of variants with zero effect (pi0; inverse of polygenicity). The SNP-based heritability was estimated from the GWAS summary statistics using LD-score regression (LDSC) (Bulik-Sullivan et al., 2015). However, pi0 is challenging to estimate from GWAS summary statistics, and so a range of pi0 values were used (0.92, 0.94, 0.96, 0.98). As a sensitivity analysis, AVENGEME analysis was also performed using the SNP-based heritability estimated by AVENGEME’s ‘estimatePolygenicModel’, using the observed pT+clump polygenic score associations.

The AVENGEME estimated AUC/*R*^2^ of polygenic scores based on LDSC and AVENGEME SNP-based heritability and a range of pi0 parameters are shown in Figure S6-S7. The figures show the observed AUC/*R*^2^ of pT+clump polygenic scores across a range of p-value thresholds, and the observed AUC/*R*^2^ of DBSLMM polygenic scores. The results show that AVENGEME estimates of AUC/*R*^2^ are not affected by the pi0 parameter when the p-value threshold (pT) is equal to 1. However, pT=1 is often not the optimal pT, particularly for less polygenic phenotypes, and therefore using the AVENGEME estimate of AUC/*R*^2^ based on a pT=1 can lead to an underestimation of AUC/*R*^2^. There are two important observations from the results.

First, the accuracy of AVENGEME estimates of AUC/*R*^2^ is influenced by LDSC SNP-based heritability estimates. For example, LDSC SNP-based heritability for MultiScler is low, leading to low estimates of AUC/*R*^2^ by AVENGEME (Table S1). However, when the SNP-based heritability of MultiScler is estimated by AVENGEME using the observed pT+clump associations, the SNP-based heritability is higher, leading to an increased AUC/*R*^2^ estimate. It is not clear which method is providing the most accurate estimate of SNP-based heritability, but the discordance between the two approaches can lead to in accurate estimates of AUC/*R*^2^.

Second, AVENGEME is designed to model pT+clump polygenic scores and is therefore not well suited to polygenic scores derived using more modern shrinkage-based polygenic scoring methods, such as DBSLMM. The results show that the estimated AUC/*R*^2^ of polygenic scores for the best pT and a pi0 parameter of 0.94 is similar to the observed AUC/*R*^2^ of the DBSLMM polygenic score. However, assuming a pi0 parameter of 0.94 does not allow for differences in polygenicity across phenotypes, leading to underestimation of AUC/*R*^2^ for low polygenicity phenotypes, and overestimation of AUC/*R*^2^ for high polygenicity outcomes (Table S3-S4).

These results indicate the AVENGEME/LDSC approach is not well suited to estimation of AUC/*R*^2^ for polygenic scores derived using approaches such as DBSLMM based on summary statistics alone.

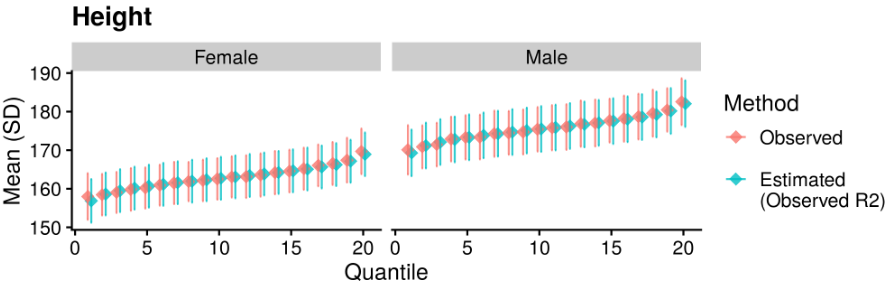

Figure S1. Validation of conversion to absolute scale using height stratified by sex. Compares observed and estimated phenotype mean and standard deviation across 20 polygenic score quantiles. Estimated values are based on the observed polygenic score R^2^ within males and females.

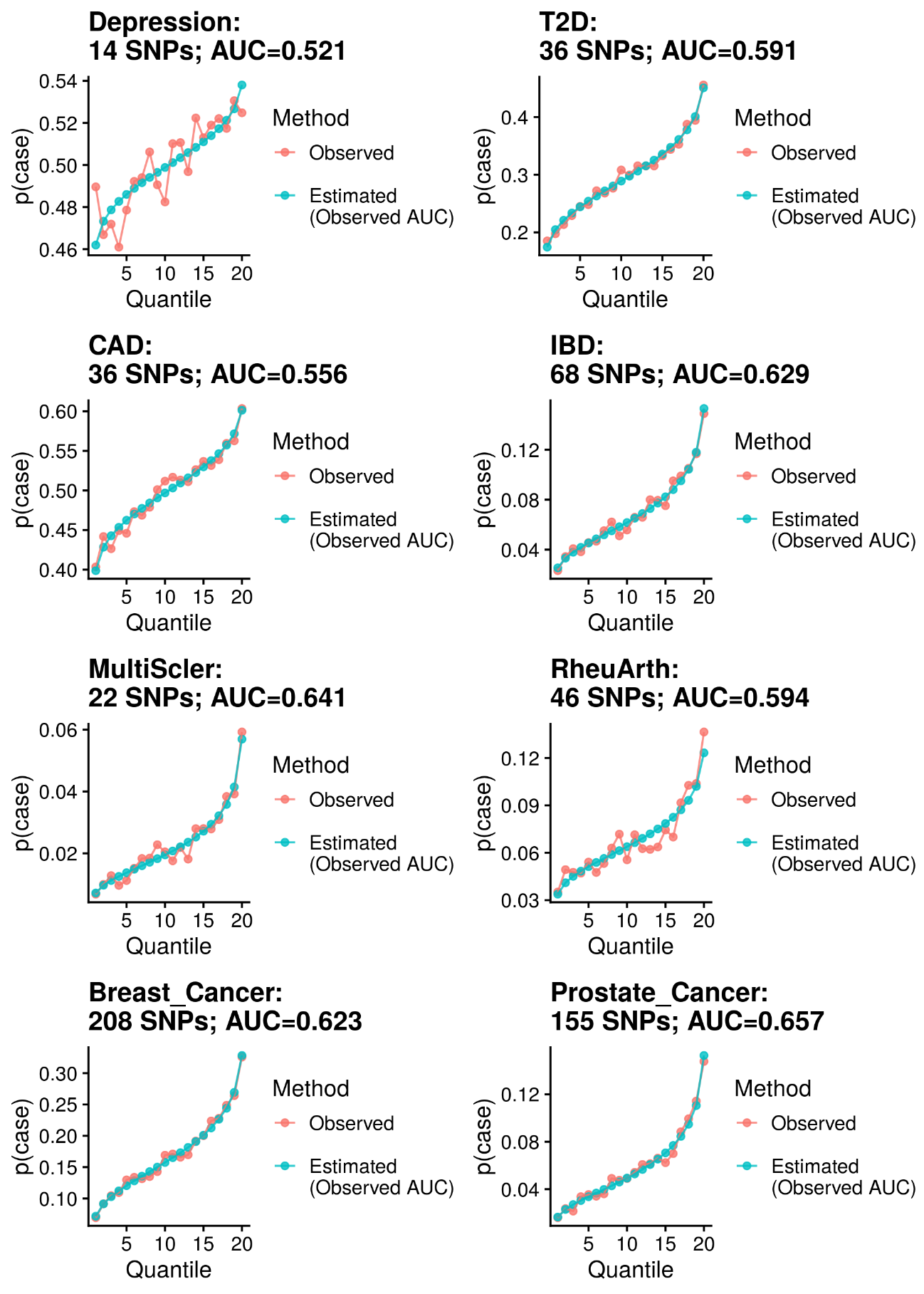

Figure S2. Comparison of observed and estimated probability of being a case across 20 pT+clump polygenic score quantiles. The pT+clump polygenic scores are derived using the most stringent p-value threshold retaining at least 5 variants. Estimated values are based on the observed polygenic score AUC. The number of SNPs considered in the polygenic score, and the AUC of the polygenic score are shown for each phenotype.

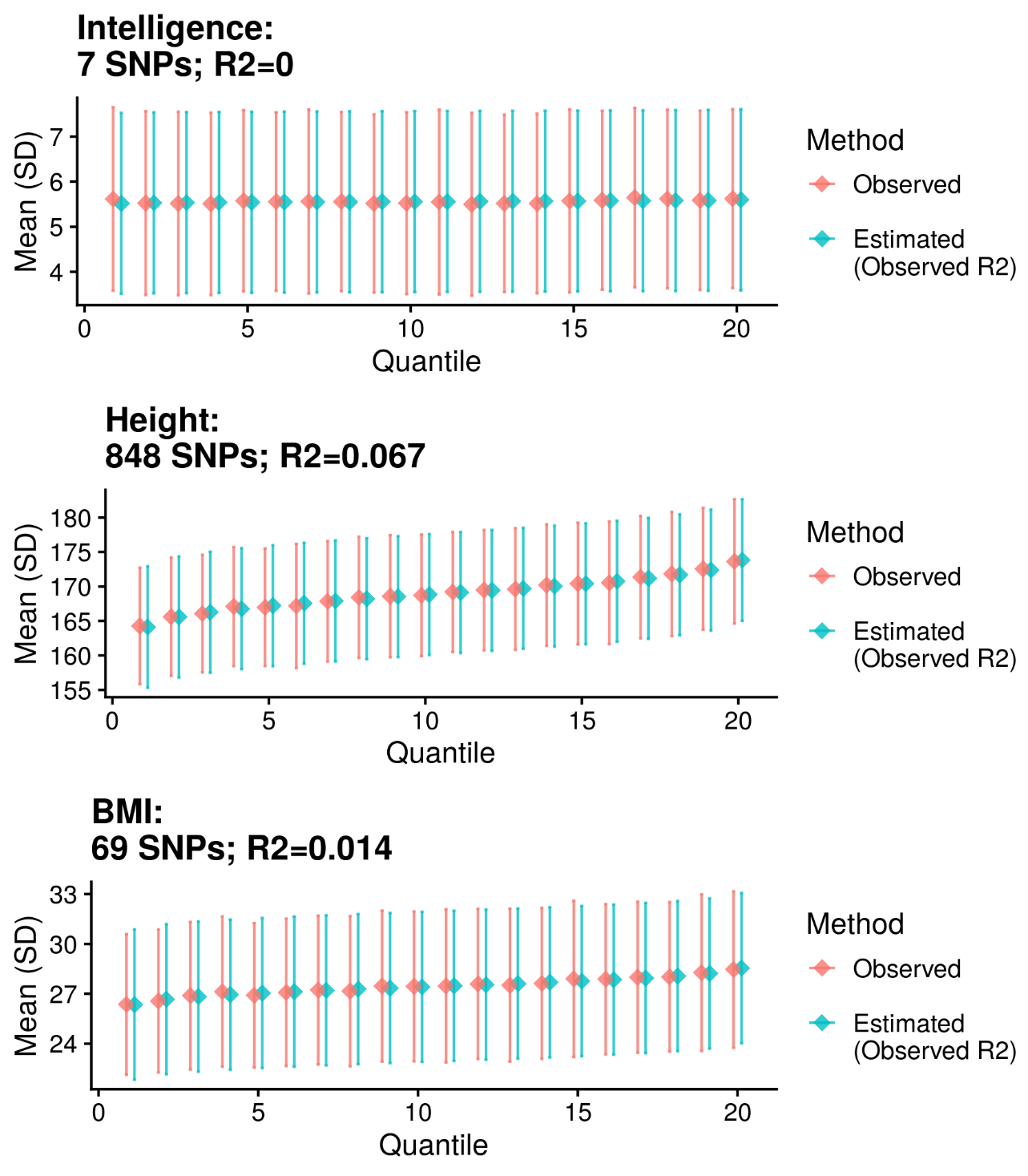

Figure S3. Comparison of observed and estimated phenotype mean and standard deviation across 20 pT+clump polygenic score quantiles. The pT+clump polygenic scores are derived using the most stringent p-value threshold retaining at least 5 variants. Estimated values are based on the observed polygenic score R^2^. The number of SNPs considered in the polygenic score, and the R^2^ of the polygenic score are shown for each phenotype.

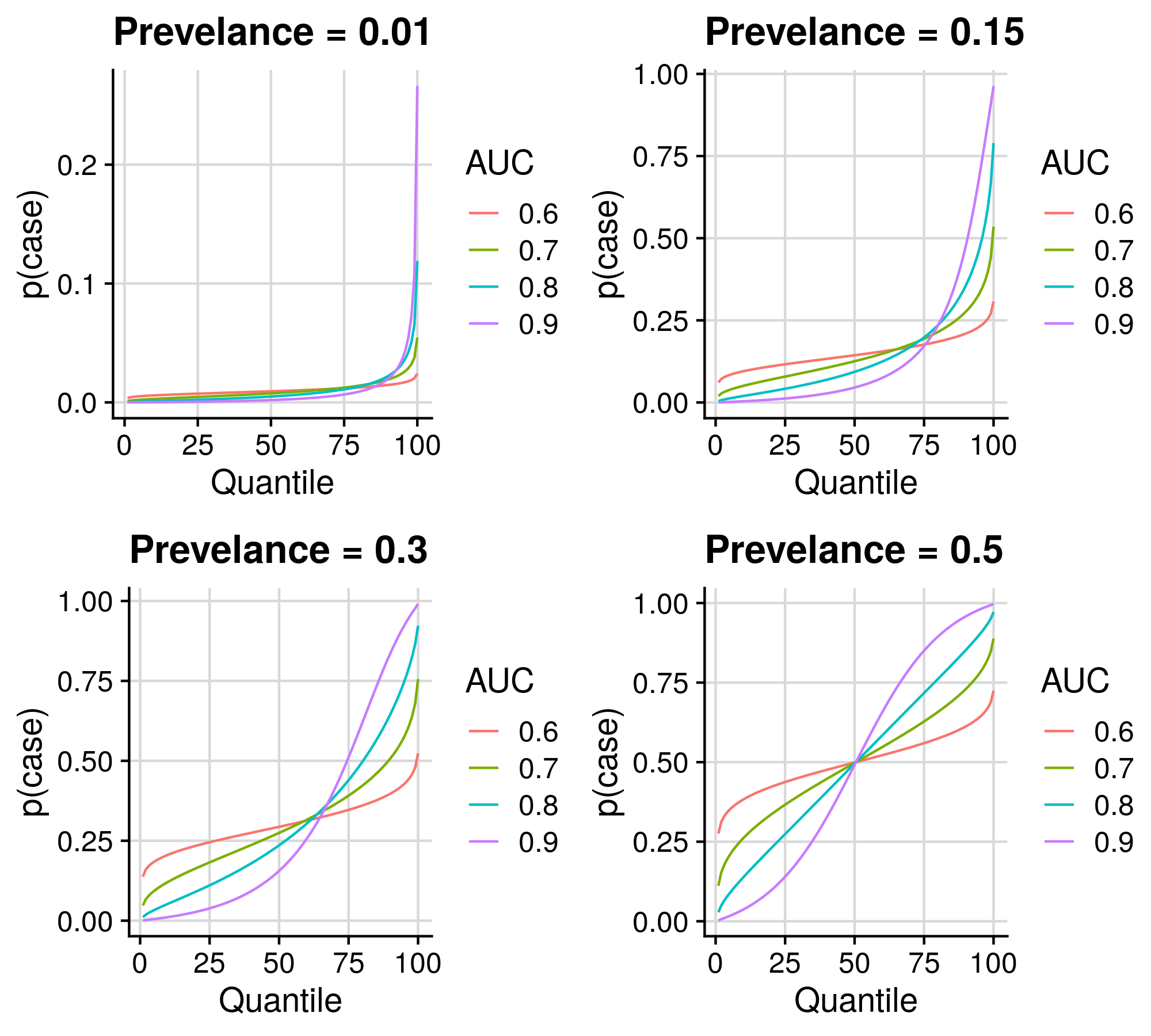

Figure S4. Absolute risk across polygenic score quantiles given a range of polygenic scores AUC and prevalence values. Y-axis shows the proportion of cases per polygenic scores quantile.

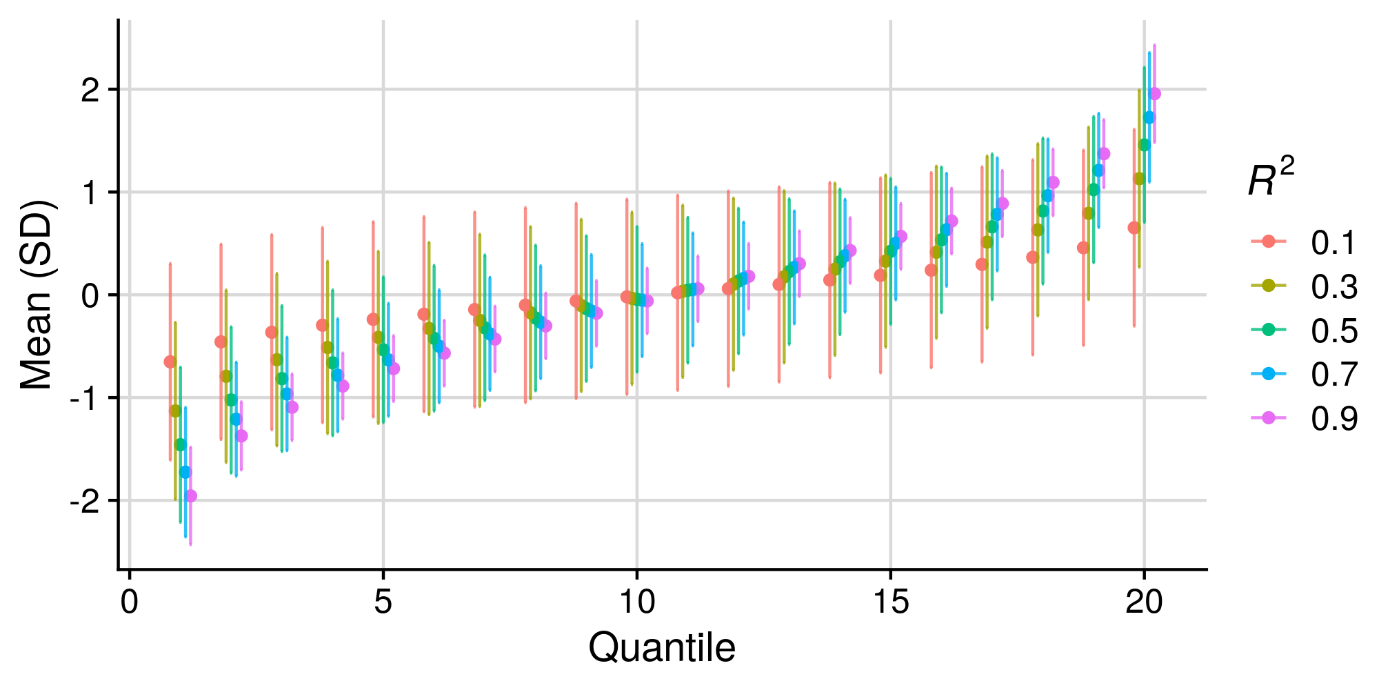

Figure S5. The phenotypic mean and SD across polygenic scores quantiles given a range of polygenic scores R^2^ values.

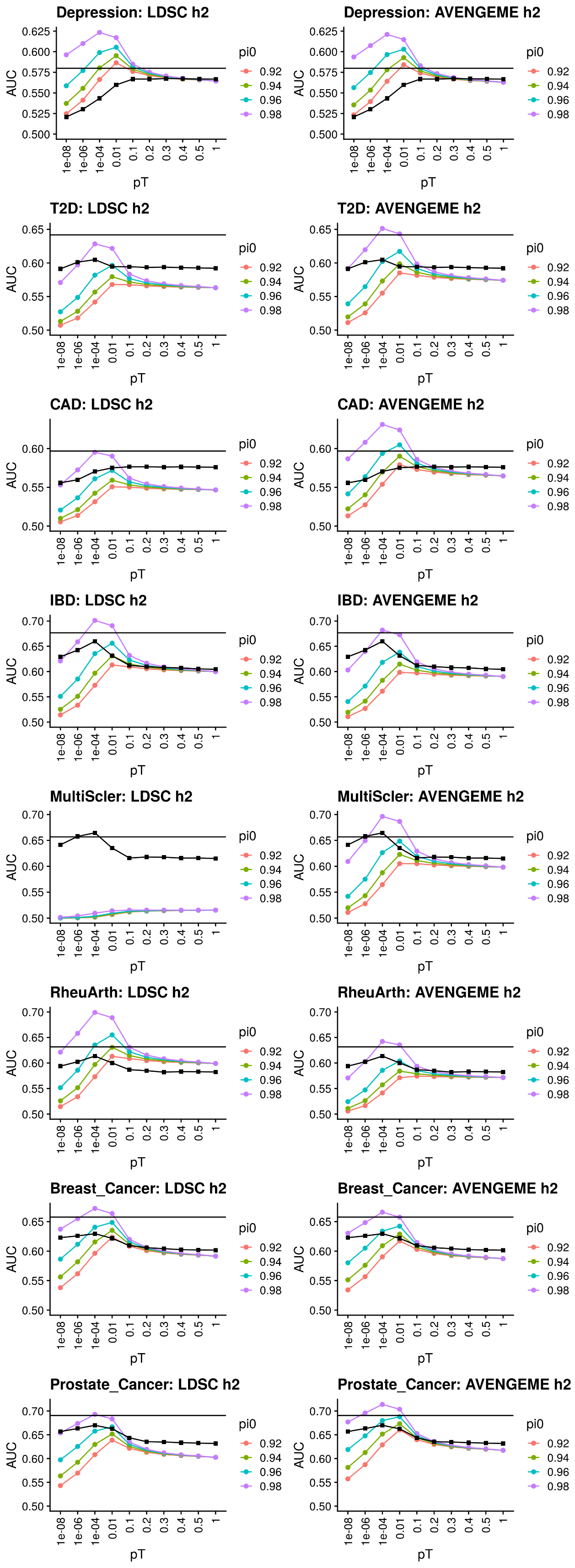

Figure S6. Part 1: Comparison of observed polygenic score AUC values and AVENGEME estimates across p-value thresholds (pT) and observed. The black points indicate the observed AUC when using polygenic scores derived using the pT+clump approach. The horizontal black line indicates the observed AUC of the polygenic score derived using the DBSLMM approach. AVENGEME estimates are provided using a range of pi0 values, indicating the proportion of variants with zero effect. On the left are AVENGEME AUC estimates when using the LDSC SNP-based heritability estimate. On the right are AVENGEME AUC estimates based on AVENGEME estimated SNP-based heritability based on observed pT+clump associations in UK Biobank.

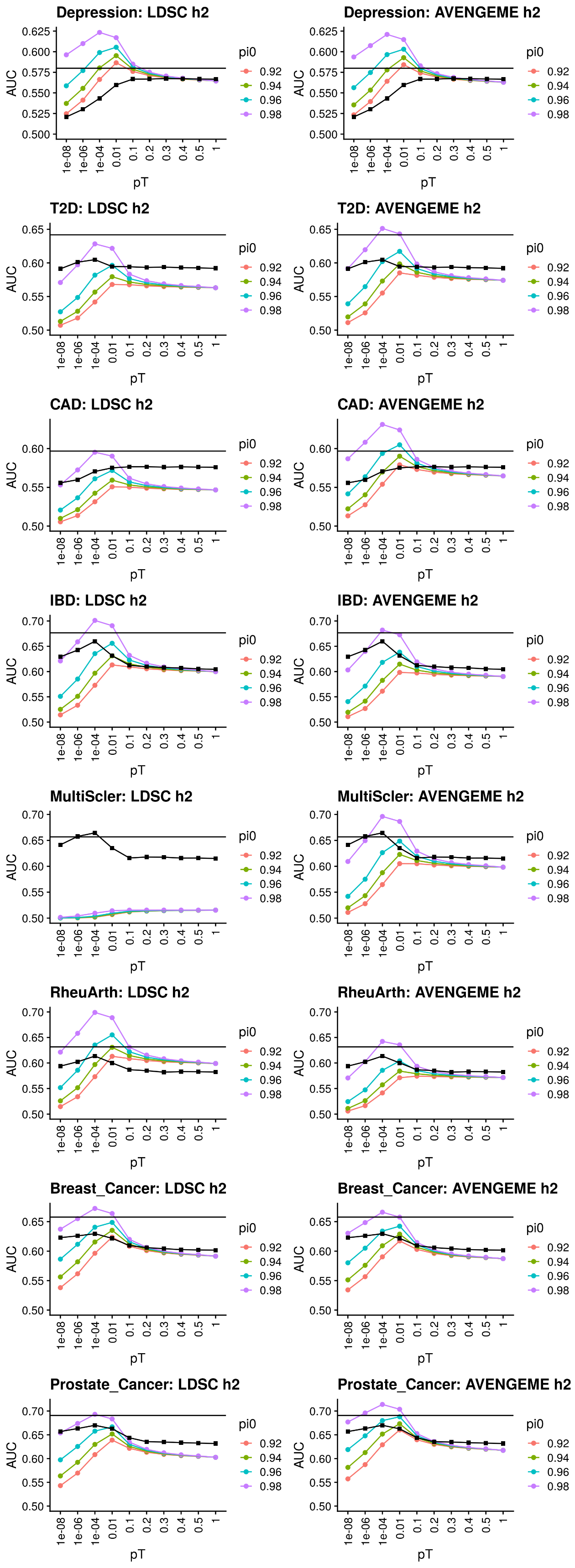

Figure S6. Part 2: Comparison of observed polygenic score AUC values and AVENGEME estimates across p-value thresholds (pT) and observed. The black points indicate the observed AUC when using polygenic scores derived using the pT+clump approach. The horizontal black line indicates the observed AUC of the polygenic score derived using the DBSLMM approach. AVENGEME estimates are provided using a range of pi0 values, indicating the proportion of variants with zero effect. On the left are AVENGEME AUC estimates when using the LDSC SNP-based heritability estimate. On the right are AVENGEME AUC estimates based on AVENGEME estimated SNP-based heritability based on observed pT+clump associations in UK Biobank.

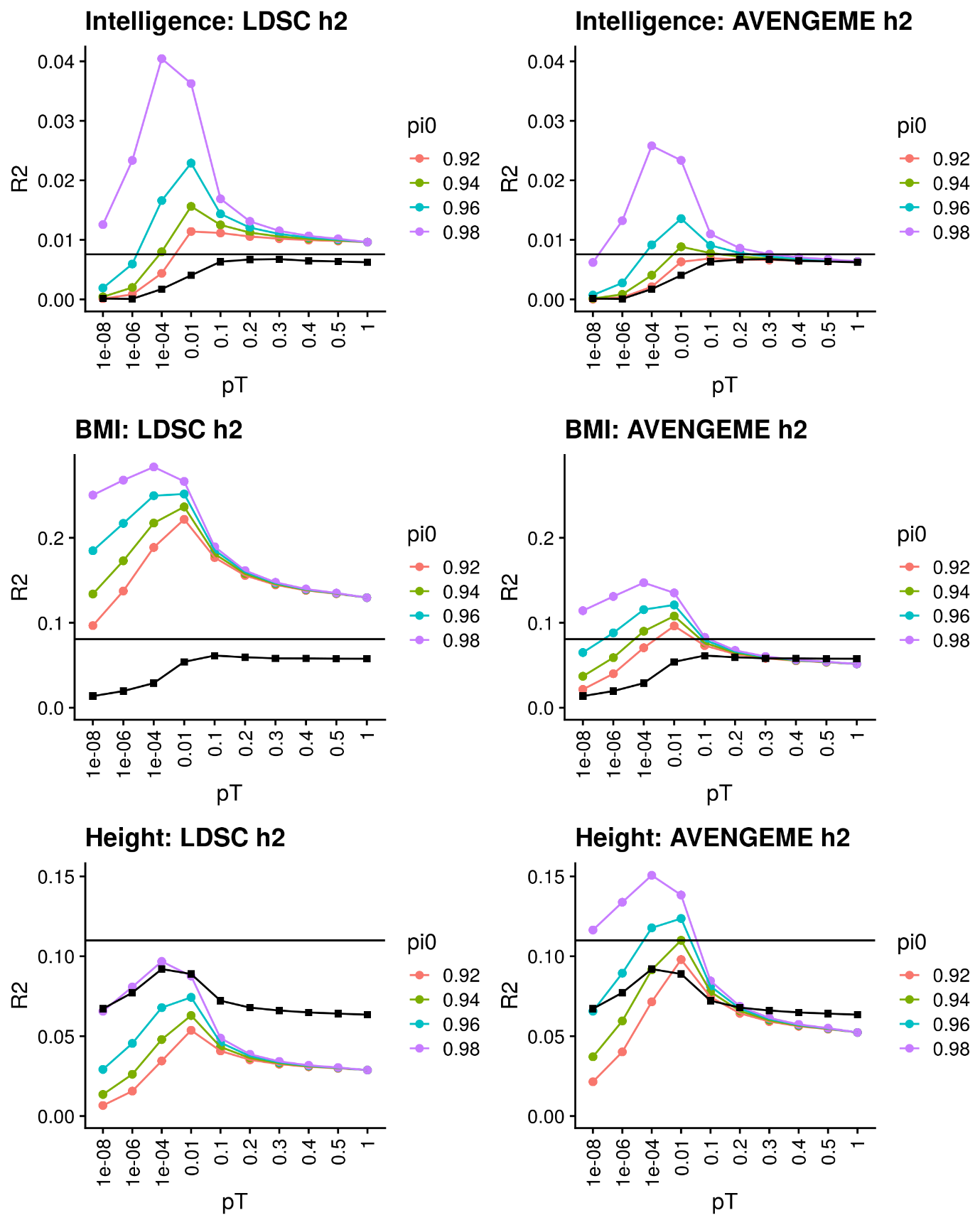

Figure S7. Comparison of observed polygenic score R^2^ values and AVENGEME estimates across p-value thresholds (pT) and observed. The black points indicate the observed R^2^ when using polygenic scores derived using the pT+clump approach. The horizontal black line indicates the observed R^2^ of the polygenic score derived using the DBSLMM approach. AVENGEME estimates are provided using a range of pi0 values, indicating the proportion of variants with zero effect. On the left are AVENGEME R^2^ estimates when using the LDSC SNP-based heritability estimate. On the right are AVENGEME R^2^ estimates based on AVENGEME estimated SNP-based heritability based on observed pT+clump associations in UK Biobank.

#### References

Bulik-Sullivan, B. K., Loh, P.-R., Finucane, H. K., Ripke, S., Yang, J., Patterson, N., … Consortium, S. W. G. of the P. G. (2015). LD Score regression distinguishes confounding from polygenicity in genome-wide association studies. *Nature Genetics*, *47*(3), 291–295.

Davis, K. A. S., Coleman, J. R. I., Adams, M., Allen, N., Breen, G., Cullen, B., … Holliday, J. (2020). Mental health in UK Biobank–development, implementation and results from an online questionnaire completed by 157 366 participants: a reanalysis. *BJPsych Open*, *6*(2).

Fürtjes, A. E., Coleman, J. R. I., Tyrrell, J., Lewis, C. M., & Hagenaars, S. P. (2020). Phenotypic Associations and Shared Genetic Etiology between Bipolar Disorder and Cardiometabolic Traits. *MedRxiv*.

Glanville, K. P., Coleman, J. R. I., Hanscombe, K. B., Euesden, J., Choi, S. W., Purves, K. L., … Lewis, C. M. (2020). Classical human leukocyte antigen alleles and C4 haplotypes are not significantly associated with depression. *Biological Psychiatry*, *87*(5), 419–430.

Palla, L., & Dudbridge, F. (2015). A fast method that uses polygenic scores to estimate the variance explained by genome-wide marker panels and the proportion of variants affecting a trait. *The American Journal of Human Genetics*, *97*(2), 250–259.

Rice, M. E., & Harris, G. T. (2005). Comparing effect sizes in follow-up studies: ROC Area, Cohen’s d, and r. *Law and Human Behavior*, *29*(5), 615.

Smith, D. J., Nicholl, B. I., Breda Cullen, D. M., Ul-Haq, Z., Evans, J., Gill, J. M. R., … Hotopf, M. (2013). Prevalence and characteristics of probable major depression and bipolar disorder within UK biobank: cross-sectional study of 172,751 participants. *PloS One*, *8*(11).

Sudlow, C., Gallacher, J., Allen, N., Beral, V., Burton, P., Danesh, J., … Landray, M. (2015). UK biobank: an open access resource for identifying the causes of a wide range of complex diseases of middle and old age. *PLoS Medicine*, *12*(3).

Wilhelm, S., & Manjunath, G. B. (2015). tmvtnorm: Truncated Multivariate Normal and Student t Distribution.
